# Geographical and temporal distribution of SARS-CoV-2 globally: An attempt to correlate case fatality rate with the circulating dominant SARS-CoV-2 clades

**DOI:** 10.1101/2021.05.25.21257434

**Authors:** Rakesh Sarkar, Mamta Chawla-Sarkar, Swagata Majumdar, Mahadeb Lo, Shiladitya Chattopadhyay

## Abstract

Uncontrolled high transmission is driving the continuous evolution of SARS-CoV-2, leading to the nonstop emergence of the new variants with varying sensitivity to the neutralizing antibodies and vaccines.Wehave analysed of 8,82,740 SARS-CoV-2 genome sequences, collected and sequenced during late December 2019 to 25 March 2021 from all across the world. The findings revealed differences in temporal and spatial distribution,and predominance of various clades/variants among six different continents.We found no clear association between the pathogenic potential of the various clades by comparing the case fatality rate (CFR) of 170 countries with the predominant SARS-CoV-2 clades in those countries, demonstrating the insignificance of the clade specific mutations on case fatality. Overall, relying on a large-scale dataset,this study illustratedthe time-basedevolution andprevalence of various clades/variantsamong different geographic regions.The study may help in designing continent specific vaccines in the future.

## 1. Introduction

The novel coronavirus disease 2019 (COVID-19) caused by severe acute respiratory syndrome coronavirus 2 (SARS-CoV-2) was first reported in late December, 2019 in the city of Wuhan, China. As of 31March, 2021, about one year since the pandemic was declared, the disease had spread to 221 countries across the world, infecting around 132 million people and causing2.8 million casualties (**https://www.worldometers**).Since its eminence, the virus has been continuously evolving by procuring new mutations which may also be a crucial factor in driving rapid transmission and governing severity of the disease. Notably, the nature of pandemic’s progression and the case fatality rate (CFR) vary among different countries. This is possibly due to difference in demographic composition of the population, the government policies to mitigate transmission of the disease and most importantly the pathogenic potential of different SARS-CoV-2 clades (Forster et al., 2020).

Phylogenetic studies, till early March, 2020, reported the emergence of three different clades of SARS-CoV-2 namely A, B and C (**Mercatelli et al., 2020**), which were subsequently named L, S and V clade respectively by the Global Initiative on Sharing All Influenza Data (GISAID) (**https://www.gisaid.org/**). With the progression of the pandemic, five more different clades (Clade G, GH, GR, GV and O according to GISAID nomenclature) having specific set of novel coexisting mutations emerged and predominated until December 2020. The G clade is characterized by the presence of four coexisting mutations 241C>T in 5’-UTR, F106F (3037C>T) in NSP3, P323L (14408C>T) in RdRp and D614G (23403A>G) in S glycoprotein; whereas the GH, GR and GV clades have additional Q57H (25563G>T) in ORF3a, RG203KR (28881GGG>CCA) in N protein, and A222V (22227C>T) in S glycoprotein respectively **(Hodcroft et al., 2020; To et al., 2020; Ceraolo et al., 2020**). The missense mutation L84S (28144T>C) in ORF8 and the silent mutation S76S in NSP4 (8782C>T) are the signature mutations of the S clade (**Brufsky et al., 2020**). The V clade is distinguished by the hallmark mutations G251V (26144G>T) in ORF3a and L37F (11083G>T) in NSP6 (**Mercatelli et al., 2020**). The first emerging L clade carries the reference alleles for all the loci defined in the G, GH, GR, GV, S, O and V clades. Genome sequences not matching the mutational criteria of above mentioned six clades are considered as the O clade (**Hodcroft et al., 2020**).

Emerging SARS-CoV-2 variants with evidence of increased transmissibility, increased severity, reduced sensitivity to neutralizing antibodies and reduced vaccine-induced protection were termed “variants of concern” (VOCs) by CDC (**https://www.cdc.gov**). At the transition of 2020-2021 four VOCs emerged, each with several mutations throughout the genome, including the S gene, and rapidly predominated globally. The VOC202012/01 (Variant of Concern, year 2020, month 12, variant 01) (also known as B.1.1.7 or 20I/501Y.V1 or GRY or UK variant) contains 17 lineage defining non-synonymous mutations, including the D614G mutation and 8 additional mutations in Spike glycoprotein: ΔH69-V70, ΔY144, N501Y, A570D, P681H, T716I, S982A, and D1118H (**Rambaut et al., 2020, Wang et al., 2021)**. The VOC B.1.351 (also known as 20H/501Y.V2 or South African variant) had 9 spike mutations in addition to D614G, including a cluster of mutations (Δ242-244&R246I) in NTD, three mutations (K417N, E484K, & N501Y) in RBD, and one mutation (A701V) near the furin cleavage site (**Tegally et al., 2020; Wang et al., 2021**). The VOC P.1 (also known as Brazilian variant or 20J/501Y.V3 or B.1.1.248) encompasses 10 spike mutations in addition to D614G, including three mutations (K417N/T, E484K, and N501Y) in the receptor-binding domain (RBD), five mutations (L18F, T20N, P26S, D138Y and R190S) in the N-terminal domain (NTD), and H655Y near the furin cleavage site (**Faria et al., 2021; Sabino et al., 2021**). The VOC P.1.427/429 (also known as California variant or 20C/S:452R) holds five mutations in addition to D614G, including three mutations (S13I; W152C; L452R) in spike protein, one in OFR1a (I4205V) and one in OFR1b (D1183Y) (**Zhang et al., 2021**).

High transmissibility or infection rate of SARS-CoV-2 had enabled the virus to acquire continuous mutations, having the potential to give enhanced fitness, that lead to the emergence of multiple clades despite being low evolutionary rate of SARS-CoV-2 (∼0.8 ×10^−3^ substitutions per site per year) compared to other RNA viruses (**Day et al., 2020; Fontanet et al., 2021**). Several studies have been attempted to uncover the relevance of gained mutations in the context of viral spread and immune invasion. Primary importance was given to spike protein mutations because this glycoprotein is involved in receptor binding and entry into the host cell, and is also the prime target for neutralizing antibodies (**Ou et al., 2020; Piccoli et al., 2020**). Several studies have claimed the role of D614G mutation of spike protein in amplified infectivity in *vitro* (**Korber et al., 2020; Hou et al., 2020**), enhanced spread in hamsters (**Hou et al., 2020**) and a mild escalation in neutralization susceptibility (**Weissman et al., 2020**), all of which were demonstrated by D614G induced more open conformation of the receptor binding domain (RBD) of the spike protein (**Yurkovetskiy et al., 2020)**.The A222V mutation of S glycoprotein, which is present within the B cell immuno-dominant epitope, has also been linked with the reinfection of SARS-CoV-2 (**Ceraolo et al., 2020**).The functional relevance of mutations acquired within the non-structural and structural proteins except spike protein remains largely undetermined. Though, recent studies have claimed the role of L37F mutation of NSP6 in asymptomatic infection based on clinical data (**Aiewsakun et al., 2020; Wang et al., 2020**). Therefore, the overall pathogenic potential and/or transmissibility of SARS-CoV-2 could be influenced by cumulative effect of all the mutations selected in different genes throughout the genome. It is still unclear whether the difference in case fatality rate and speed of disease transmission among different countries are due to discrepancy in the pathogenic potential of different clades or variants.

In the present study, we provided a snapshot of geographical and temporal distribution of different clades all across the world to give a better understanding of the gradual emergence and predominance of different clades/variants over time. We also attempted to evaluate the pathogenic potential of different clades by correlating the case fatality rate of 170 countries with the SARS-CoV-2 clades that were prevalent in those countries.

## 2. Materials and Methods

### 2.1. Sequence retrieval and analysis

In total, we analyzed 8,82,740SARS-CoV-2 genome sequences, that were collected during late December, 2019 to 25 March, 2021 from all across the world, available at the global initiative of sharing all influenza data (GISAID) **(https://www.gisaid.org/)** (**Shu et al., 2017**). Distribution of different clades of SARS-CoV-2among various continents and associated countries were analyzed by using “Location” and “Clade” specific filters offered by GISAID database, whereas temporal distribution of different clades/variants was analyzed by using “Location”, “Collection Date” and “Clade/Variant” specific filters. First isolated SARS-CoV-2 strains of different clades/variants from six different continents were identified by initially sorting sequences using “Location” and “Clade/variant” specific filters and then analysing the first collection date.

### 2.2. CFR calculation

We have accessed WHO site (**https://covid19.who.int/**) on 25 March, 2021 to compile the total number of reported COVID cases and confirmed COVID deaths in various countries. Then, case fatality rate (CFR) of each country was calculated by using the formula: (Total number of COVID deaths/Total number of confirmed COVID cases) X 100.

## 3. Results

### 3.1. Geographical root monitoring and temporal distribution of SARS-CoV-2 globally

In the present study, we have analyzed 8,82,740 genome sequences of SARS-CoV-2 deposited in GISAID from 170 countries of six different continents during late December 2019 to March 2021. Most of the genomes were deposited from Europe (560187 samples, 63.46%) followed by North America (235458 samples, 26.67%), Asia (48012 samples, 5.44%), Oceania (19063 samples, 2.16%), Africa (10557 samples, 1.20%) and South America (9463 samples, 1.07%) (**Figure 1A, Supplementary Table 1**). Frequency distribution of different clades/variants showed the worldwide predominance of the B.1.1.7 (214443, 24.29%) followed by the GH (183066, 20.74%), GR (167474, 18.97%), GV (138595, 15.70%)and G (117588, 13.32%) clades, which together constitute around 93% of the total sequences. The frequencies of the L, O, S, V, B.1.351, P.1, B.1.427/B1.429 and B.1.525 clades range from 0.10-1.96% (**Figure 1B, Supplementary Table 1**). Geographical root monitoring based on first collection date and month wise frequency distribution of different clades revealed the first detection of the L clade (the first emerged clade) and the S clade in December 2019 at Wuhan of China. The V clade was first collected from China in January 2020. These three clades (L, S and V) surged in March 2020 and subsequently declined. In January 2020, the four clades (G, GH, GR and GV) emerged in Europe. The G clade was first detected in Germany whereas the GH, GR and GV clades were first detected in Netherlands. The G, GH and GR clades remained dominant from March 2020 to August 2020. The GV clade suddenly surged in September, 2020 and dominated over G, GH and GR clades until December, 2020. Though, we found the first appearance of the B.1.1.7 or UK variant in Spain in February 2020, B.1.1.7 continued to surge from October 2020 and dominated worldwide from January 2021 to March 2021. Initial emergence of the B.1.427/429 or California variant was seen in Mexico in July, 2020 followed by continuous increase in frequency from November 2020. After initial detection in South Africa in October 2020, the B.1.351 (South African variant) gradually spread out with the progression of time. The Brazilian variant (P.1) and the Nigerian variant (B.1.525) were first seen in Brazil and Nigeria respectively in December 2020, however no significant escalation in their frequencies were witnessed in the subsequent months (**Table 1, Table 2, Figure 1C, Supplementary Table 2**). Overall, we observed worldwide predominance of the GH clade in March 2020, the GR clade from April 2020 to September 2020, the GV clade from October 2020 to December 2020, and the UK variant from January 2021 to March 2021 (**Figure 1C**).

**Table 1:**
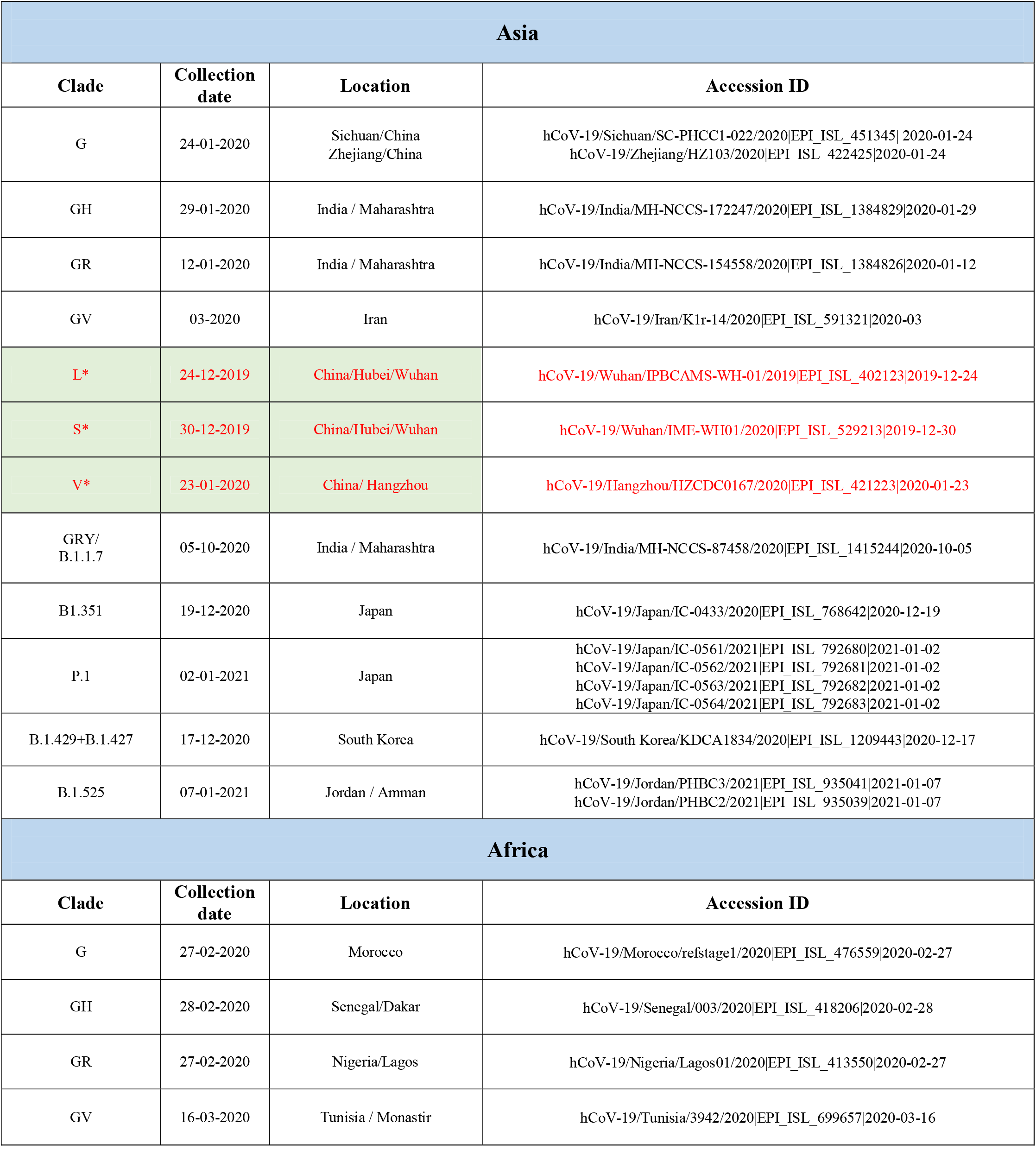
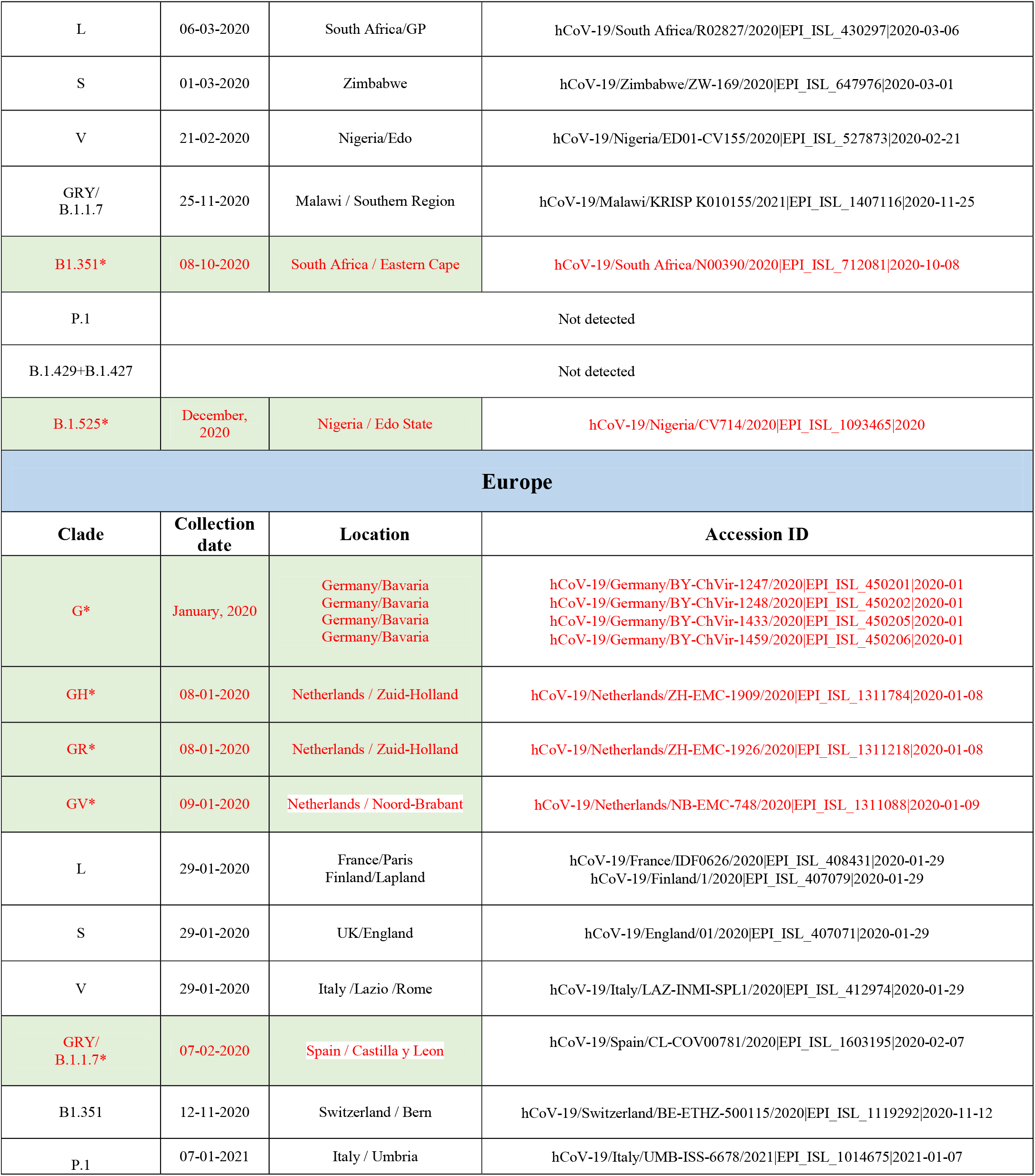
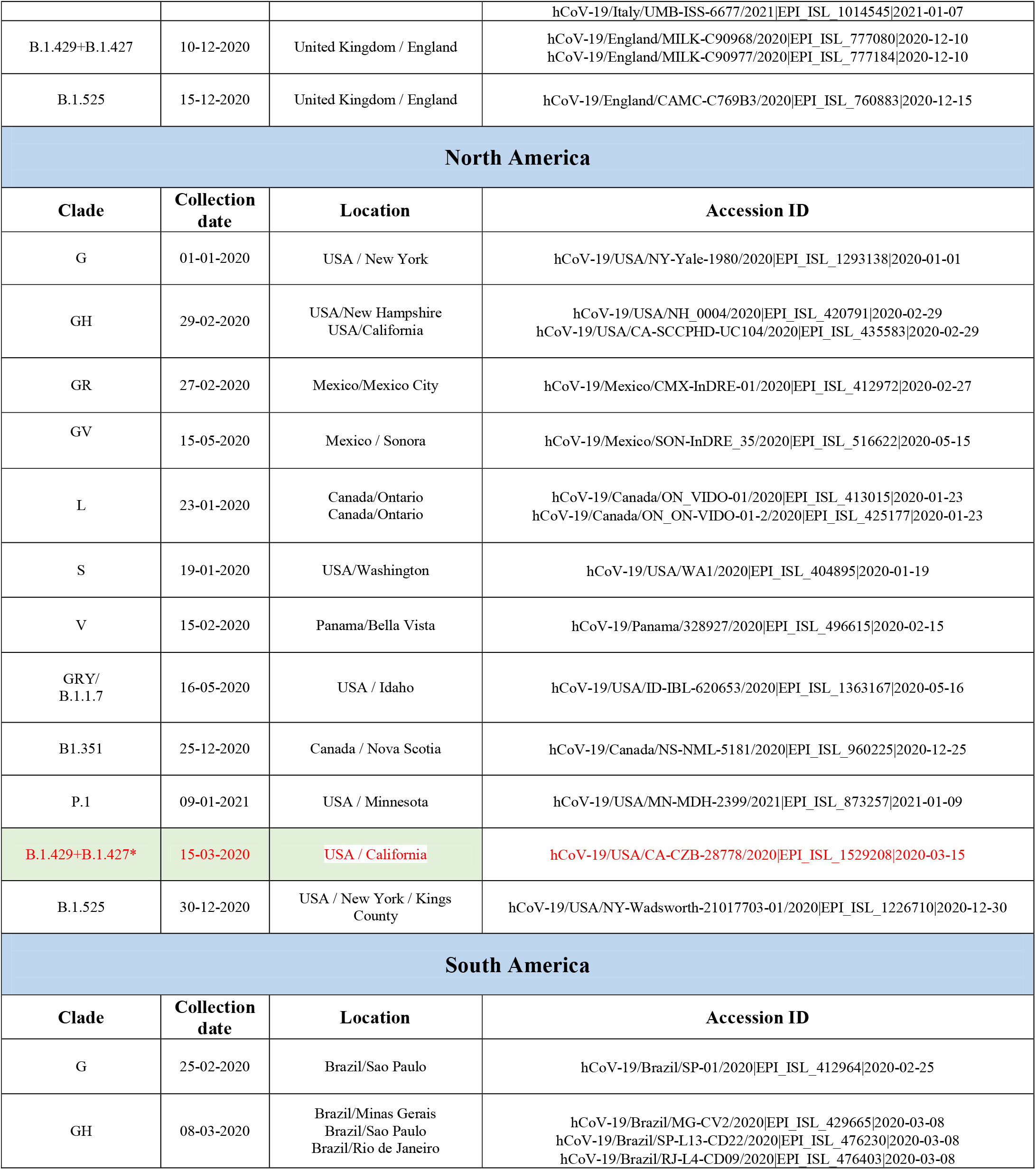
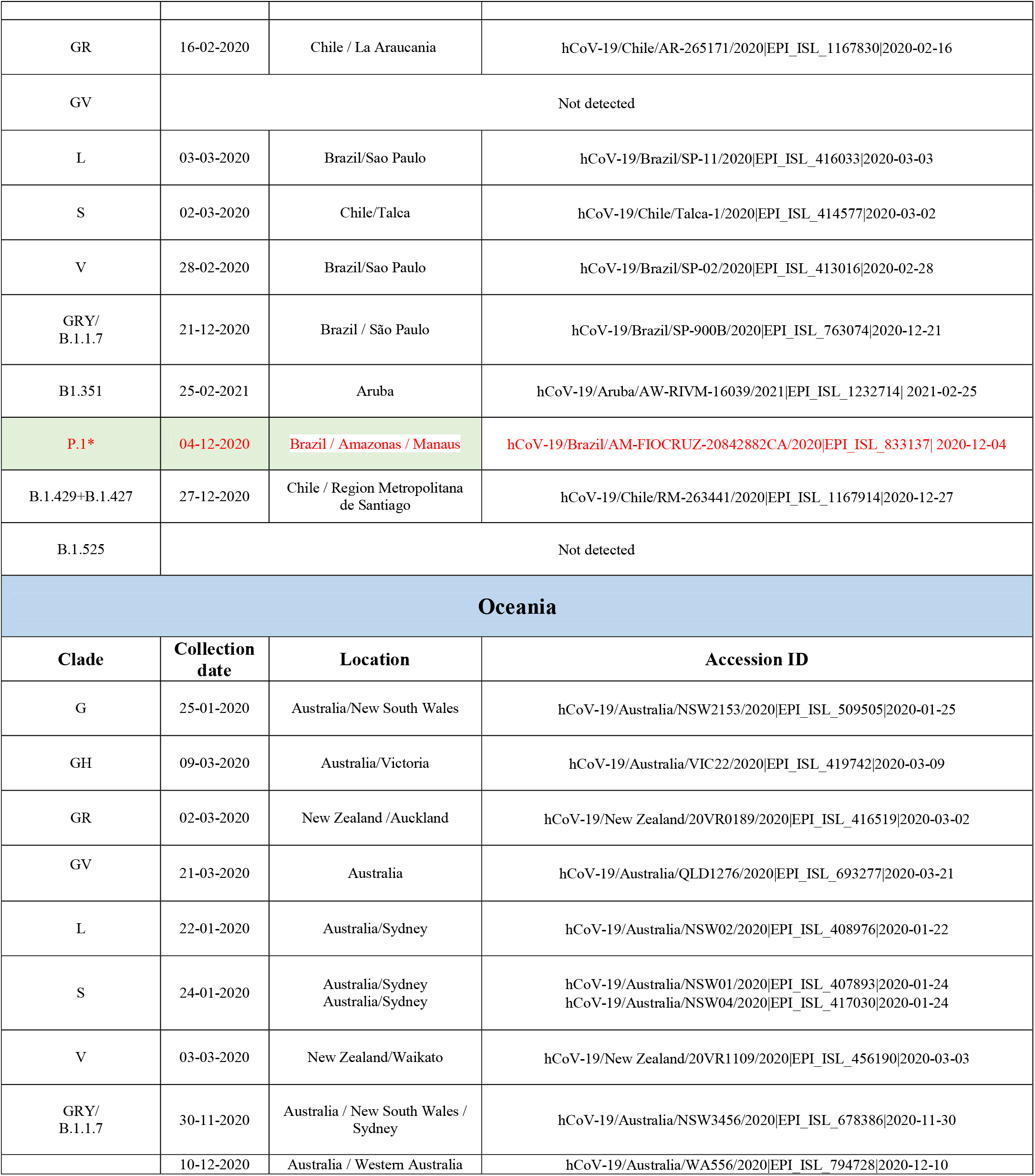
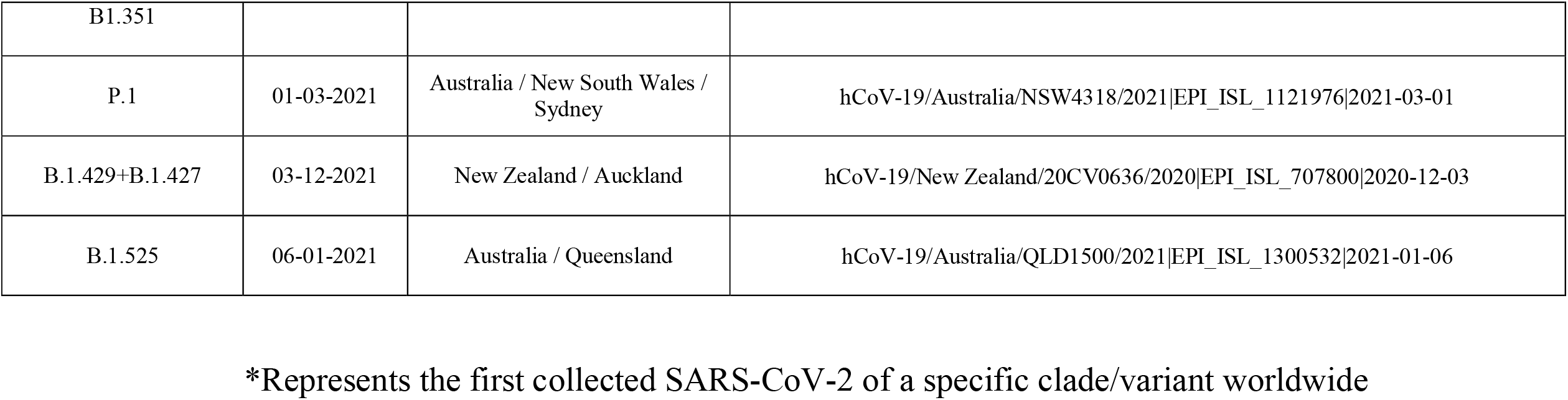
Details of the first collected SARS-CoV-2 of twelve different clades from six differentcontinents

**Table 2:** Temporal dominance of different clades/variants of SARS-CoV-2 in six different continents

|  | 2020 |  |  |  |  |  |  |  |  |  |  |  | 2021 |  |  |  |
| --- | --- | --- | --- | --- | --- | --- | --- | --- | --- | --- | --- | --- | --- | --- | --- | --- |
|  | Jan | Feb | Mar | Apr | May | Jun | Jul | Aug | Sep | Oct | Nov | Dec | Jan | Feb | Mar |  |
| Worldwide |  |  | G,GH,GR |  |  |  |  |  | GV |  |  |  | B.1.1.7 |  |  |  |
| Asia |  |  | GR |  |  |  |  |  |  |  |  |  |  | B.1.1.7 |  |  |
| Africa |  |  | G |  |  | GR |  |  |  |  | B.1.351 |  |  |  | B.1.525 |  |
| North America |  |  | GH |  |  |  |  |  |  |  |  |  |  |  |  |  |
| South America |  |  | GR |  |  |  |  |  |  |  |  |  |  |  |  | G |
| Europe |  |  | GR |  |  |  |  |  | GV |  |  |  | B.1.1.7 |  |  |  |
| Oceania |  |  | GH |  |  | GR |  |  |  |  | GH |  |  |  |  |  |

**Figure 1:**
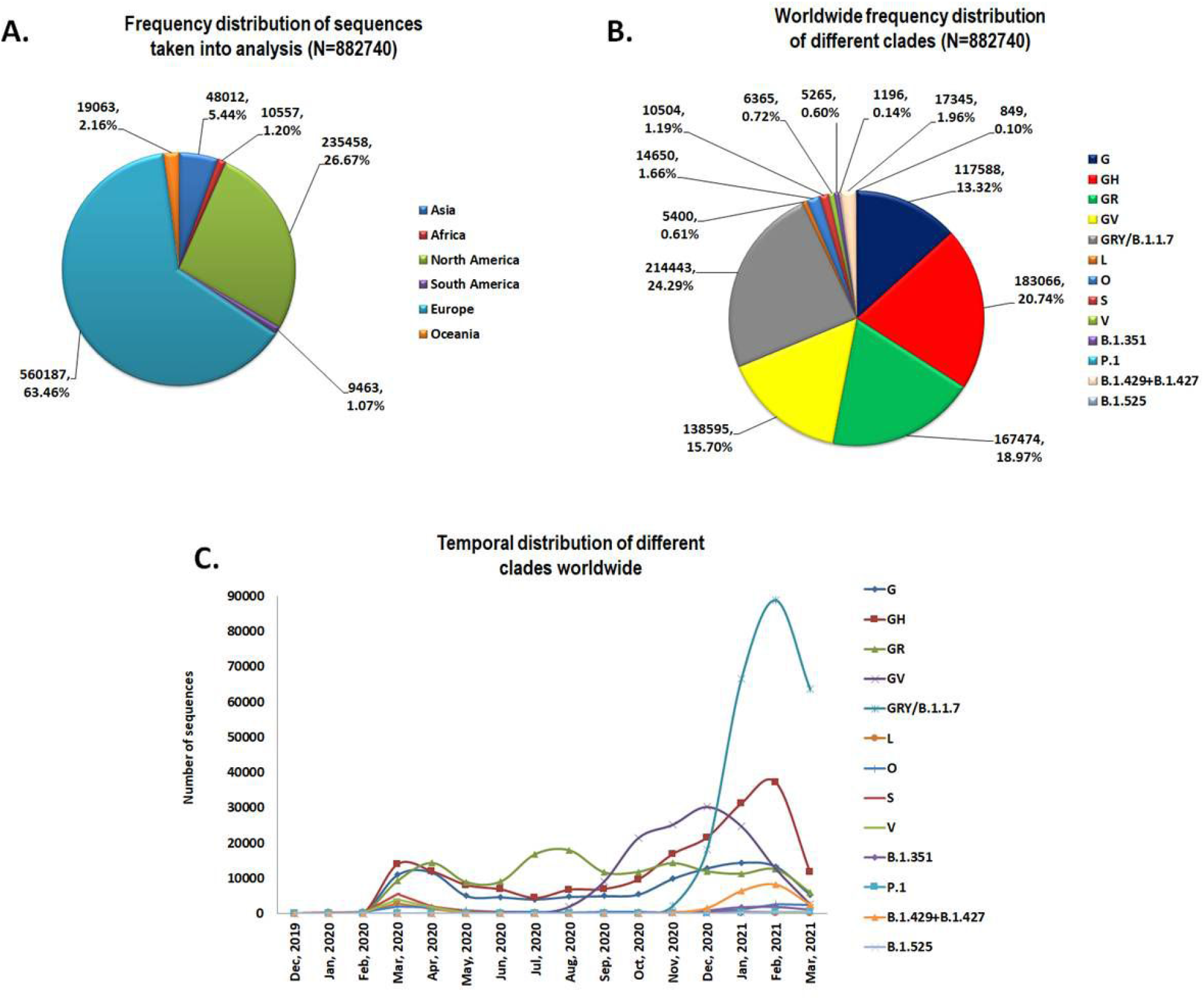
(A) Frequency distribution of SARS-CoV-2 sequences taken into analysis from six different continents. (B) Global frequency distribution of different SARS-CoV-2 clades. (C) Worldwide temporal distribution of different SARS-CoV-2 clades from December, 2019 to March, 2021.

### 3.2. Temporal distribution of SARS-CoV-2 in Asia

The frequency distribution of various clades in Asia is different from the worldwide scenario. Asia was mostly predominated by the GR clade (28544, 59.45%) followed by GH (7681, 16.00%), G (3155, 6.57%), O (3871, 8.06%), B.1.1.7 (1630, 3.39%), S (1377, 2.87%) and L (1039, 2.16%). The frequency of the each of GV, V, B.1.351, P.1, B.1.427/B1.429 and B.1.525 clades was less than 1% (**Figure 2A, Supplementary Table 3**). Temporal distribution presented that Asia has been consistently dominated by the GR clade, first observed in India among all the Asian countries in January 2020, since March 2020 to January 2021. In December 2020, the UK variant made its first appearance in India among Asian countries, and in February 2021, it became the dominant variant in Asia. The presence of the UK variant was detected in 24 of the 38 Asian countries studied. India, Israel, Japan, Jordan, Philippines, Singapore and South Korea all had considerably high incidence of UK variant. The South African variant and the California variant were first observed in Japan and South Korea respectively among Asian countries in December 2020, however, their occurrence remained very low throughout Asia in the following months, with the exception of the South African variant in Israel and the California variant in South Korea. In Asia, the Brazilian and Nigerian variants were first noticed in Japan and Jordan, respectively, and these mostly predominated in Japan. (**Figure 2B, Table 1, Table 2, Supplementary Figure 4**).

**Figure 2:**
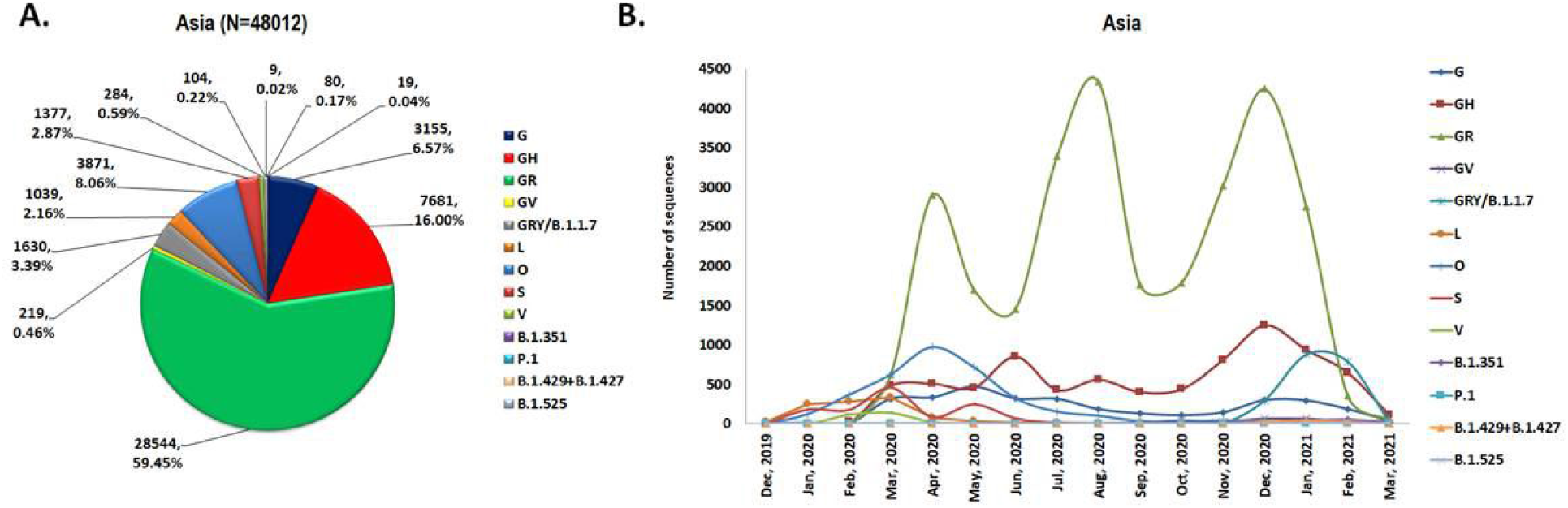
(A) Frequency distribution of different SARS-CoV-2 clades in Asia. (B)Temporal distribution of different SARS-CoV-2 clades from December, 2019 to March, 2021 in Asia.

### 3.3. Temporal distribution of SARS-CoV-2 in Africa

Overall, Africa was predominated by the GR clade (3377, 31.99%) followed by G (2716, 25.73%), B.1.351 (2222, 21.05%), GH (995, 9.43%), S (490, 4.64%), B.1.1.7 (258, 2.44%), O (247, 2.34%) and B.1.525 (116, 1.10%). The frequency of the each of GV, L and V clades was less than 1%. The Brazilian and California variants were not found in Asia among all the genome sequences available at GISAID till 25 March, 2021 (**Figure 3A, Supplementary Figure 3**). Though, the three clades G, GH, and GRwere first detected in Africa in the last week of February 2020, the G clade dominated the continent for the next three months (March 2020-May 2020). Incidence of the GR clade gradually surged from March and became the leading clade in Africa since June 2020 to October 2020, with highest frequency in July. Frequency of the GH clade remained consistently low from February 2020 to March 2021, with a pick in January 2021. After first appearance in South Africa in October 2020, the B.1.351 dominated Africa since November 2020 to February 2021. B.1.525 was first detected in Nigeria in December 2020, and by March 2021, it had established itself as the dominant strain in Africa (**Figure 3B, Table 1, Table 2, Supplementary Figure 4**).

**Figure 3:**
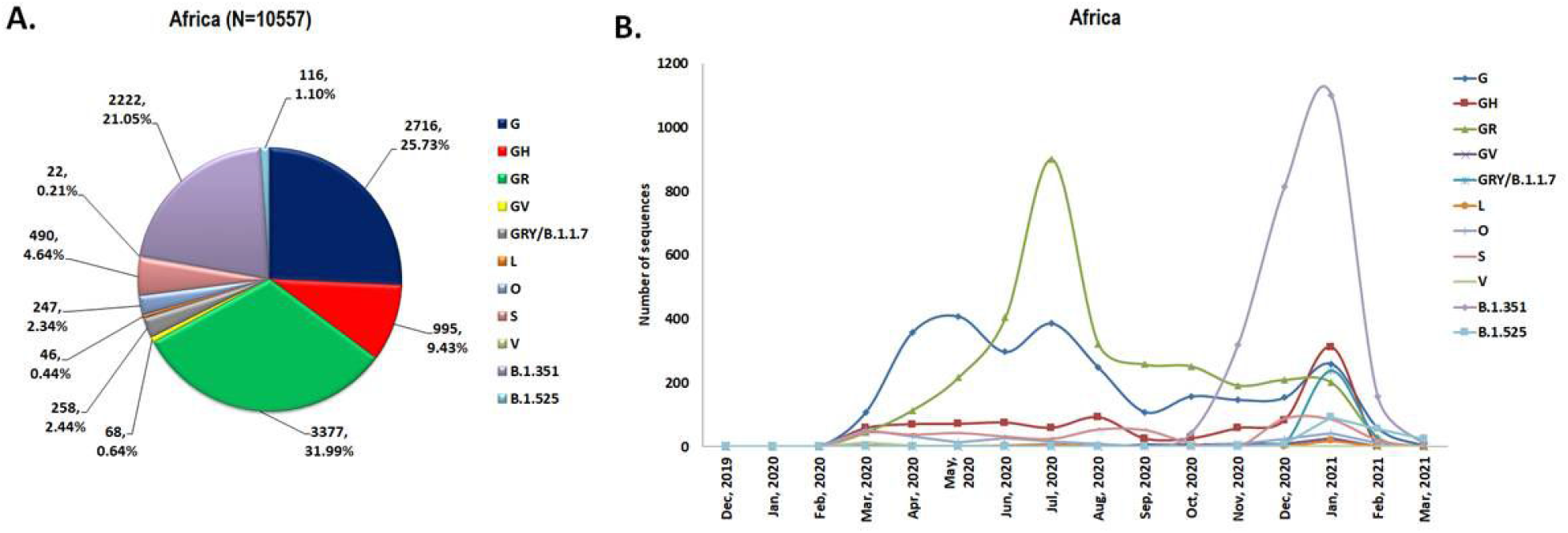
(A) Frequency distribution of different SARS-CoV-2 clades in Africa. (B)Temporal distribution of different SARS-CoV-2 clades from December, 2019 to March, 2021 in Africa.

### 3.4. Temporal distribution of SARS-CoV-2 in North America

Altogether, North America was largely predominated by the GH clade (128115, 54.41%) followed by G (43647, 18.54%), GR (30264, 12.85%), B.1.427/429 (17144, 7.28%), B.1.1.7 (7626, 3.24%) and S (4679, 1.99%) clade. The frequency of the each of GV, L, O, V, B.1.351, P.1 and B.1.525 clades was less than 1% (**Figure 4A, Supplementary Figure 3**). Temporal distribution showed that the GH clade, which first seen in USA in February 2020, consistently predominated North America since March 2020 to March 2021. Incidence of the G clade (first appeared USA in January 2020) and the GR clade (first appeared Mexico in February 2020) were comparably low to the GH clade but constantly present throughout the all months since March 2020. The California variant (B.1.427/429) and the UK variant (B.1.1.7) were first detected in California/USA in March 2020 and Idaho/USA in May 2020 respectively, but their frequencies began to rise in November 2020. The South African variant and the Brazilian variant were first observed in Nova Scotia/Canada in December 2020 and in Minnesota/USA in January 2021 respectively, and their prevalence increased in the following months (**Figure 4B, Table 1, Table 2, Supplementary Table 4**).

**Figure 4:**
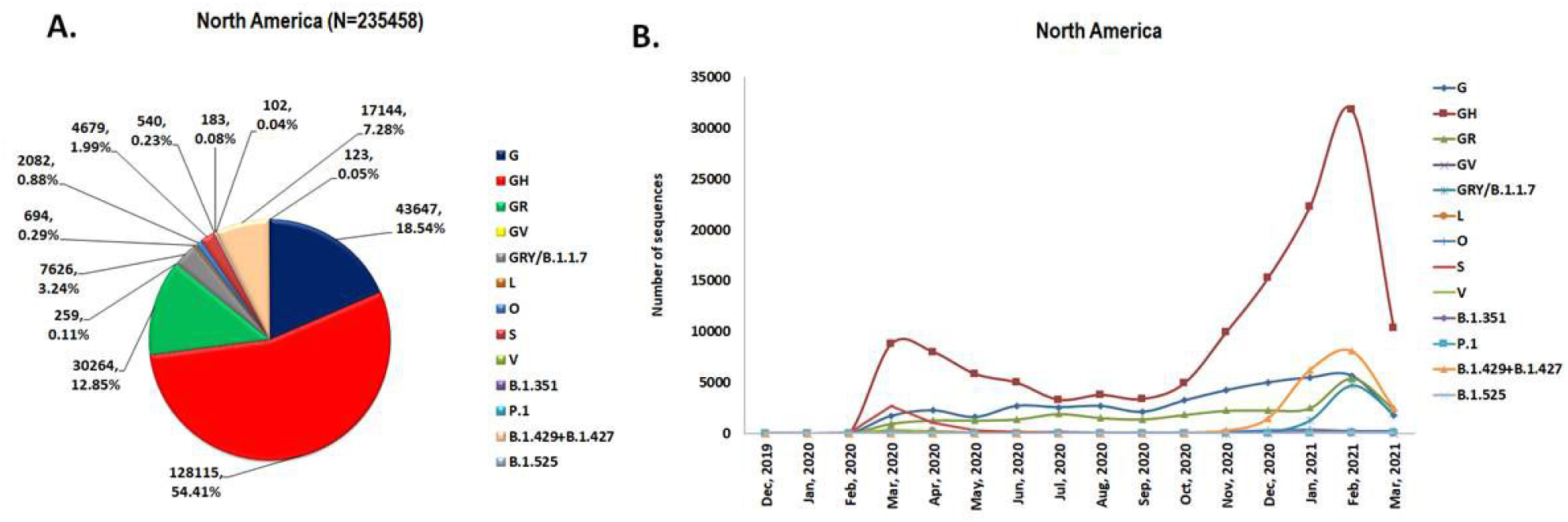
(A) Frequency distribution of different SARS-CoV-2 clades in North America. (B)Temporal distribution of different SARS-CoV-2 clades from December, 2019 to March, 2021 in North America.

### 3.5. Temporal distribution of SARS-CoV-2 in South America

South America was mostly dominated by the GR clade (6087, 64.32%) followed by GH (1228, 12.98%), G (1011, 10.68%), P.1 (496, 5.24%), B.1.1.7 (263, 2.78%) and O (169, 1.79%). The frequency of the each of GV, L, S, V, B.1.351, B.1.427/429 clades was less than 1%. The Nigerian variant (B.1.525) was not seen in South America (**Figure 5A, Supplementary Table 3**). Spatial and temporal distribution showed the predominance of the GR clade in South America since its first detection in Chile in February 2020 to February 2021, with highest occurrence in April 2020. The G and GH clades, which were first detected in Brazil in February and March 2020 respectively, peaked in frequencies in April and May of 2020, and then steadily declined in frequency until November 2020. The incidence of the G clade has risen suddenly since December, 2020 and became the predominant clade in South America in March, 2021. The Brazilian variant was first isolated from Brazil in December, 2020 and peaked in frequencies in January, 2021 and declined subsequently. Since its first emergence in December 2020, the UK variant has gradually increased in frequency (**Figure 5B, Table 1, Table 2, Supplementary Table 4**).

**Figure 5:**
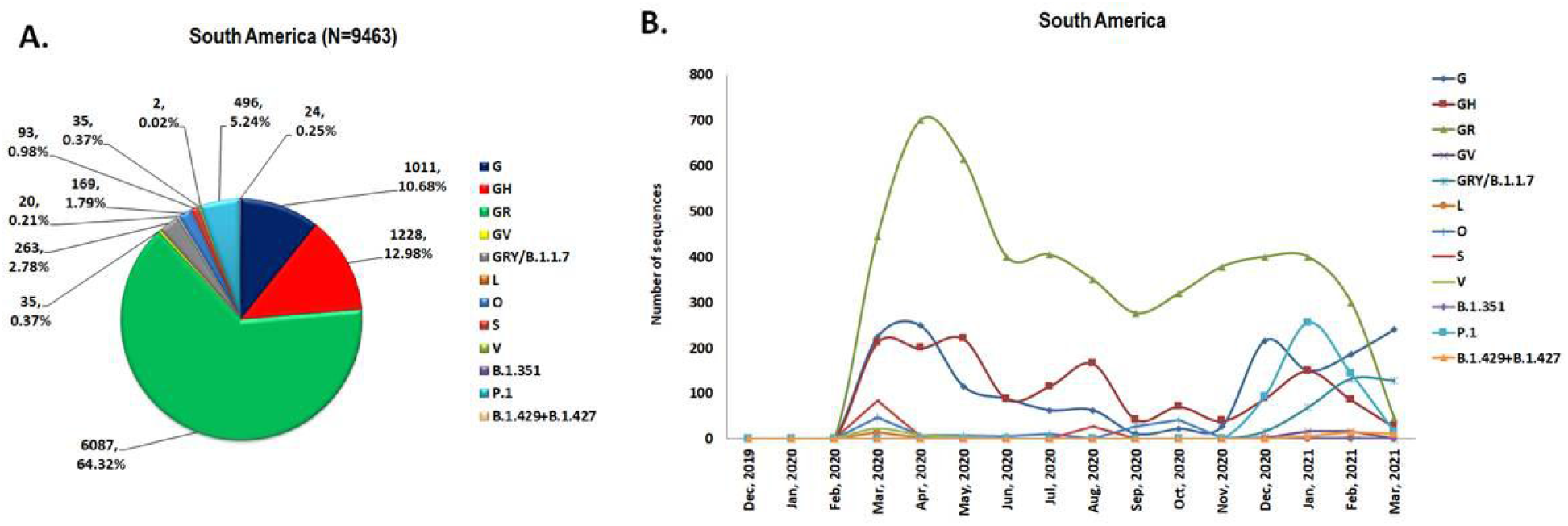
(A) Frequency distribution of different SARS-CoV-2 clades in South America. (B)Temporal distribution of different SARS-CoV-2 clades from December, 2019 to March, 2021 in South America.

### 3.6. Temporal distribution of SARS-CoV-2 in Europe

Geographical distribution of different clades showed Europe was largely predominated by B.1.1.7 clade (204407, 36.49%) followed by the GV (137937, 24.62%), GR (85301, 15.23%), G (65836, 11.75%), GH (43340, 7.74%) and O (7769, 1.39%). The frequency of the each of L (3530, 0.63%), S (2973, 0.53%), V (5154, 0.92%), B.1.351 (2702, 0.48%), P.1 (548, 0.10%), B.1.427/429 (69, 0.01%) and B.1.525 (585, 0.10%) clades was less than 1% (**Figure 6A, Supplementary Figure 3**). Though, four clades G, GH, GR and GV were first isolated from Europe in January 2020 with respect to the global scenario, the GR clade predominated Europe until August 2020. The GV clade surged in frequency since August 2020 and dominated Europe from September 2020 to December 2020 with a steady rise in prevalence. The UK variant was first collected from Spain in February 2020, however, started to surge in frequency from December 2020 and dominated Europe from January 2021 to March 2021. The South African variant (First detected in Switzerland in November 2020), the Brazilian variant (First detected in Italy in January 2021), the California variant (First detected in England in December 2020) and the Nigerian variant (First detected in England in December 2020) remained very low in frequencies after their first detection (**Figure 6B, Table 1, Table 2, Supplementary Table 4**).

**Figure 6:**
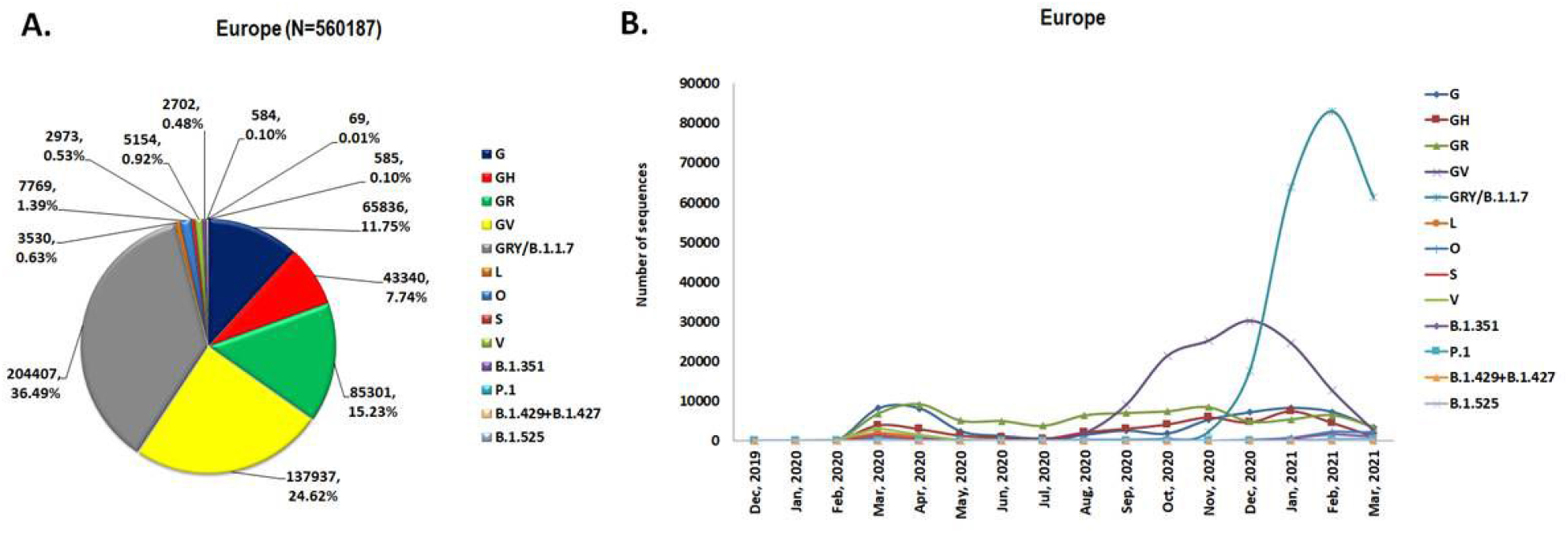
(A) Frequency distribution of different SARS-CoV-2 clades in Europe. (B)Temporal distribution of different SARS-CoV-2 clades from December, 2019 to March, 2021 in Europe.

### 3.7. Temporal distribution of SARS-CoV-2 in Oceania

Oceania was largely predominated by the GR clade (13901, 72.92%) followed by the GH (1707, 8.95%), G (1223, 6.42%), S (892, 4.68%), O (512, 2.69%), V (330, 1.73%) and B.1.1.7 (259, 1.36%). Each of the GV, L, B.1.351, P.1, B.1.427/429 and B.1.525 clades had a frequency of less than 1% (**Figure 7A, Supplementary Table 3**). In Oceania, the G clade was first detected in Australia and surged in March 2020. The GH and GR clades were first detected in March 2020 in Australia and New Zealand respectively. The GH clade dominated Oceania from March 2020 to May 2020 and also from November 2020 to March 2021, whereas GR clade prevailed during June to October 2020. The UK variant was first detected in Australia in November 2020 and became the second most prevalent clade after GH in 2021. No significant surge in frequencies of the South African variant (First detected in Western Australia in December 2020), the Brazilian variant (First detected in Sydney/Australia in March 2021), the California variant (First detected in Auckland/New Zealand in December 2020), and the Nigeria variant (First detected in Australia in January 2021) was noticed after their first detection (**Figure 7B, Table 1, Table 2, Supplementary Table 4**).

**Figure 7:**
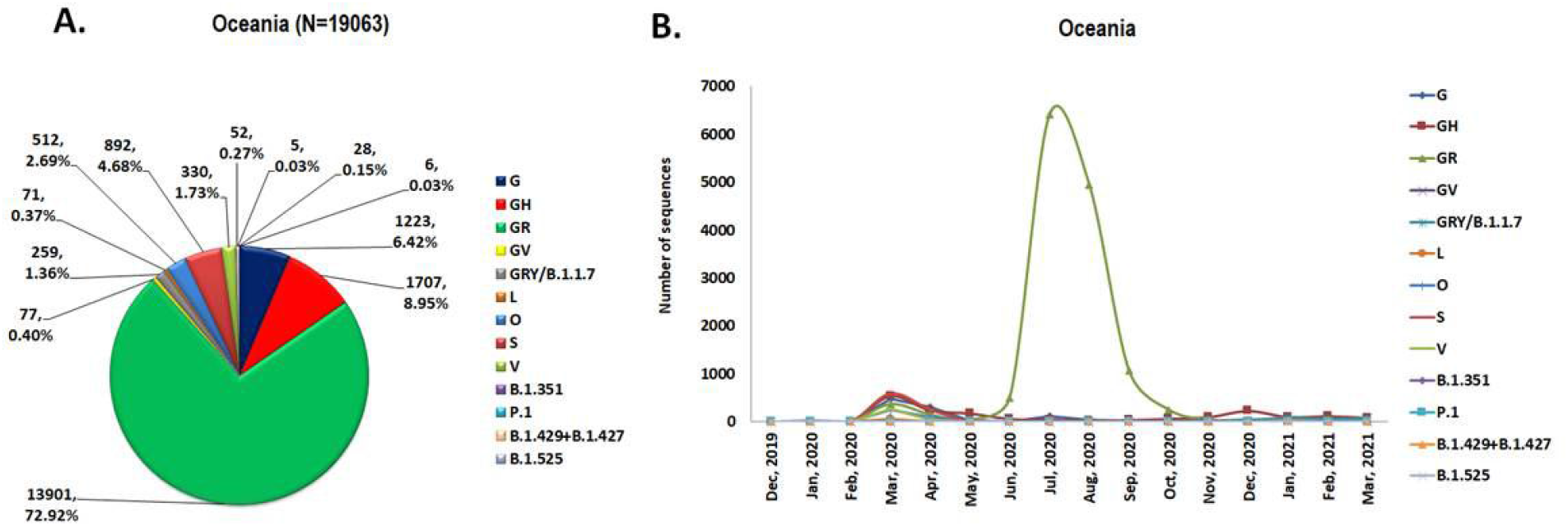
(A) Frequency distribution of different SARS-CoV-2 clades in Oceania. (B)Temporal distribution of different SARS-CoV-2 clades from December, 2019 to March, 2021 in Oceania.

### 3.8. The impact of the circulating dominant clade over a country’s CFR

We have estimated the CFR of 170 countries all around the world, including 38 countries from Asia, 39 countries from Africa, 22 countries from North America, 18 countries from South America, 48 countries from Europe and 5 countries from Oceania. Though, the global CFR was estimated to be 2.19, we observed a wide range of CFR across countries, ranging from 0.00 to 9.06. Among 170 countries, 7 countries had CFR of >4, 17 countries had CFR between 3 to 4, 31 countries had CFR between 2 to 3, 71 countries had CFR between 1-2 and 44 countries had CFR between 0 to 1 (**Table 3, Supplementary Table 5)**. Estimation of the CFR by continents showed that Asia had the lowest average CFR (1.58) among the six continents, with 38 countries having CFRranging from 0.00 to 4.72. Africa had the highest average CFR (2.64) among the continents, with CFRs ranging from 0.55 to 5.94 in 39 countries.The average CFRs for Europe, North America, Oceania, and South America were 2.20 (48 countries, CFR range: 0.15-3.88), 2.30 (22 countries, CFR range: 0.00-9.06), 2.52 (5 countries, CFR range: 1.00-3.10), and 2.60 (18 countries, CFR range: 0.00-5.15), respectively (**Table 3, Supplementary Table 6**).

**Table 3:** Case fatality rate (CFR) of different continents

| **Continent** | **Total confirmed cases** | **Total confirmed deaths** | **CFR** | **CFR range among countries** | **Number of countries** |
| --- | --- | --- | --- | --- | --- |
| **Asia** | 24527049 | 387471 | 1.58 | 0.00-4.72 | 38 |
| **Europe** | 42773531 | 942005 | 2.20 | 0.15-3.88 | 48 |
| **North America** | 34390178 | 791741 | 2.30 | 0.00-9.06 | 22 |
| **Oceania** | 44790 | 1127 | 2.52 | 1.00-3.10 | 5 |
| **South America** | 21043537 | 547963 | 2.60 | 0.00-5.15 | 18 |
| **Africa** | 4070145 | 107440 | 2.64 | 0.55-5.94 | 39 |
| **Worldwide** | **126849230** | **2777747** | **2.19** | **0.00-9.06** | **170** |

To evaluate the effect of the dominant clade on a country’s CFR, we further looked at the prevalence of various clades in each of the 170 countries. We noticed that most of the countries were dominated by the G clade (32 countries, CFR range among countries: 0.08-4.52), GH clade (42 countries, CFR range among countries: 0.00-4.39), GR clade (42 countries, CFR range among countries: 0.33-9.06) and O clade (14 countries, CFR range among countries: 0.00-3.35). UK variant and South African variant were found to be prevalent among 13 countries (CFR range: 0.40-3.87) and 6 countries (CFR range: 0.80-4.13), respectively. Surprisingly, we did not notice any strong correlation between the dominant clade and country’s CFR; each of the clades was responsible for varying levels of CFRs (low to high) among different countries where they were predominant (**Table 4, Supplementary Table 7**).

**Table 4:** CFR range of countries predominated by specific clade(s)

| **Predominant clade** | **CRF range among countries** | **Number of countries** |
| --- | --- | --- |
| G | 0.08-4.52 | 31 |
| GH | 0.00-4.39 | 42 |
| GR | 0.33-9.06 | 42 |
| GV | 0.47-3.06 | 8 |
| L | 4.72 | 1 |
| S | 0.55-3.87 | 8 |
| O | 0.00-3.35 | 14 |
| G+GH | 0.12-3.52 | 2 |
| GH+GR | 1.09-2.55 | 2 |
| GH+GR+O | 1.64 | 1 |
| GRY/B.1.1.7 | 0.40-3.87 | 13 |
| B.1.351 | 0.80-4.13 | 6 |

## 4. Discussion

All viruses are immensely potent in adapting to new hosts (**Sanjuan et al., 2016**). Genotypic diversity among viruses occurs due to the accumulating mutations that might lead to various phenotypic changes in them. The continuous replacement of the branches in the tree of life (clade) is the key characteristic of biodiversity (**Silvestro et al., 2015**). The viral biodiversity could be addressed by phylogenetic studies by analyzing the nucleotide/ amino acid homology (**Das et al., 2021**). Clades are the monophyletic groups of organisms that share a common ancestor along with all its lineal descendants. Various viral clades are generated through the process of evolutionary pressure on host immunogenics (**Tyor et al., 2013**). The study of the viral clades, which share sequence similarity and are descendants of a common ancestor can throw light on the evolutionary process. Different clades depict different extent of disease manifestation and pathological conditions. The host immunological determinants of disease severity are maximally gene-regulated (**Kumar et al., 2020**). Evolution of viral genome facilitates it to evade the host immunity at times (**DeDiego et al., 2008; Li et al., 2020**). Therefore, understanding the plethora of mutations in the viral genome leading to the clade-diversification could determine the importance of regulatory variation in evolution. Thorough study of the SARS-CoV-2 genomic mutations and characterizing them based on the clades from which they are derived can reveal the actual evolutionary history of the virus.

The proclivity of disease severity depends upon the type of circulating virus clade in a particular geographical region and it causes different case fatality rates across different countries. Therefore, elucidation of the SARS-CoV-2 clades with temporal and spatial differences can underscore the evolutionary crosstalk amongst them. The process of natural selection maximally determines the dynamics of an emerging mutation. The variants resulting due to mutations that impart a competitive and selection advantage in context to viral replication, transmission, or evasion of immune response will upsurge and eventually out-compete all other circulating variants. Though, other potential factors like chronic infection among immuno-compromised subjects, change of host-reservoirs, chance-events, faulty proof-reading mechanisms in the virus could be potential drivers of viral evolution. Based on the large-scale genome dataset of SARS-CoV-2, our present study depicted the global as well as continent wise emergence and prevalence of various clades/ variants as the COVID-19 pandemic progressed from its beginning in December 2019 to March 2021.

Cryo EM analyses by some researchers reveal that the N501Y mutation found among the preponderant SARS-CoV-2 UK variant (B.1.1.7) highly increases viral infectivity (**Zhu et al., 2021**). This is at par to the observation of our study where the total frequency distribution showed the global predominance of the UK variant (24.3%), currently. The UK variant possesses the characteristic feature of high transmission ability, contributing to an upsurge in the number of positive cases and hospitalizations worldwide. This is imposing an immense pressure on the health care system, especially during the second wave of the pandemic. Epidemiological surveillances and computational modelling underscore that the SARS-CoV-2 UK variant spreads 56% faster than pre-existing variants, resulting in higher nasopharyngeal viral titer compared to the prototype strain (**Davies et al., 2021a**). Studies estimate a 35% (12–64%) increased risk of death associated with this highly transmissible UK variant, indicating that it can cause serious disease manifestation, though there is no confirmatory evidence of this variant causing severity among children (**Davies et al., 2021b; Brookman et al., 2021)**. It is not evident how this B.1.1.7 achieved prominence, although the unusual genetic divergence of the B.1.1.7 lineage may have resulted, at least in part, from the evolution of the virus in an individual with chronic infection (**Rambaut et al., 2020**). Surveillance studies revealed the upsurge in SARS-CoV-2 clades harbouring the D614G mutation with the predominance of the clade GV during the last three months, which is consistent to our results. Our results underscored the temporal preponderance of mostly the GR clade, followed by others like GH, GR, GV and UK variant. As per Hamed et. al., 2021, among all the currently circulating SARS-CoV-2 clades, GR predominated worldwide, subsequently followed by GV and GH clades. Severe or deceased COVID-19 patients were more susceptible to the GH and GR clades, while the asymptomatic/ mildly symptomatic individuals revealed a higher frequency of G and GV clades (**Hamed et al., 2021**).

However, it will be worth noting that due to the recent outbreak of pandemic COVID-19, people around the world are trying by every means to reach the origin, to get some ways of prevention and therapeutic pathways. Constant global monitoring of the SARS-CoV-2 clades and variants will help in tracking viral evolution. G and G-derived clades had been reported from North America and Asia in March 2020 and are currently the predominating viral subpopulation, globally (**Mercatelli et al., 2020)**. Other in silico studies also reported the diversity of SARS-CoV-2 infection in ten Asian countries which focused on the prevalence of clades such as G, GH, GR, L, S, O, and V (**Sengupta et al., 2021**). The geographical distribution of Nextstrain and GISAID clades revealed that clade 19B/S prevailed rarely, except in Spain and Kazakhstan, while the frequencies of the rest of the clades varied across the other countries in the WHO European Region (**Alm et al., 2020**). Even the Eastern Mediterranean Region countries reported the dominance of G, GH and GR clades (**Omais et al., 2021**). Through this study, we observed an upsurge of G clade strains along with its derivatives like the GH and GR clades; while clades L and V eventually diminished. Across the United States and Spain, clade S, though declined, nevertheless can be seen sporadically.

During the first wave of the pandemic in 2020, a study revealed that the clade with variants ORF1ab L3606F, A4489V, S2015R, T2016K, and N P13L is common to Singapore which had been attributed to the low case fatality rate of Singapore (**Koyama et al., 2020**). Another study analyzed SARS-CoV-2 genomic sequences from Eastern Mediterranean Region countries and revealed no association between increased CFR and the G, GH and GR clades (**Omais et al., 2021**). A study from Washington also showed that no significant difference exists in the rates of hospitalization or death due to the predominating two clades in that region. Major risk factors for hospitalization and death for either major clade of virus include patient age and comorbid conditions(**Nakamichi et al., 2021**). In consistent with the studies by Omais et al., 2021 and Nakamichi et al., 2021, our current analysis has not found any correlation among the prevalence of a particular clade and its association to high CFR in that particular geographical area. Few clades and variants might have high infectivity, but studies in terms to their lethality are limited to date. Therefore, the differences in case fatality rate among different countries could be regulated by several factors including comorbidity risk and demographic, economic and political factors (**Sorci et al., 2020**).

In order to design the preventive measures and therapeutic approaches, tracing the evolutionary and phylogenetic history of SARS-CoV-2 is the need of the hour. Dissecting out the viral genomic diversity, clinical manifestations in conjunction with epidemiological surveillance data will certainly prove beneficial in chalking out avenues to overcome the disease burden.SARS-CoV-2 is continuing to mutate and evolve, leading to the emergence of differential genomic sequences, with higher infectivity, lethality along with clinical and pharmacological repercussions.To date, limited studies have been done to elucidate and compare the genomic, molecular and phenotypic differences between the various circulating clades of SARS-CoV-2.The current study has shed enough light on the clade-wise temporal and spatial analyses across all the 6 continents, globally. This study underscores the dynamicity of the different viral clades, which can lead to personalized/ customized prevention and treatment strategies for each geographical region depending on the circulating clade.

## Data Availability

All sequence data analyzed were freely available at the global initiative of sharing all influenza data (GISAID) (https://www.gisaid.org/)

## Acknowledgement

We would like to acknowledge all the scientists, researchers and laboratory staffs for their valued contribution in SARS-CoV-2 genome sequencing and deposition in GISAID. We would also like to applaud GISAID consortium for allowing us the open access to the deposited SARS-CoV-2 sequences.

## Author contributions

RS and M-CS designed the study, collected data, performed data analysis and wrote the paper. SW, ML and AB performed data analysis. SD provided vital guidance on the scientific content. SC conceived the study, supervised data analysis, provided scientific inputs and edited the draft manuscript.

