## Supplemental Tables for "Geographical and temporal distribution of SARS-CoV-2 globally: An attempt to correlate case fatality rate with the circulating dominant SARS-CoV-2 clades"

| Continents | G | GH | GR | GV | GRY/B.1.1.7 | L | O | S | V | B.1.351 | P.1 | B.1.429+B.1.427 | B.1.525 | Total |
|---|---|---|---|---|---|---|---|---|---|---|---|---|---|---|
| Asia | 3155 | 7681 | 28544 | 219 | 1630 | 1039 | 3871 | 1377 | 284 | 104 | 9 | 80 | 19 | 48012 |
| Africa | 2716 | 995 | 3377 | 68 | 258 | 46 | 247 | 490 | 22 | 2222 | 0 | 0 | 116 | 10557 |
| North America | 43647 | 128115 | 30264 | 259 | 7626 | 694 | 2082 | 4679 | 540 | 183 | 102 | 17144 | 123 | 235458 |
| South America | 1011 | 1228 | 6087 | 35 | 263 | 20 | 169 | 93 | 35 | 2 | 496 | 24 | 0 | 9463 |
| Europe | 65836 | 43340 | 85301 | 137937 | 204407 | 3530 | 7769 | 2973 | 5154 | 2702 | 584 | 69 | 585 | 560187 |
| Oceania | 1223 | 1707 | 13901 | 77 | 259 | 71 | 512 | 892 | 330 | 52 | 5 | 28 | 6 | 19063 |
| **Total** | **117588** | **183066** | **167474** | **138595** | **214443** | **5400** | **14650** | **10504** | **6365** | **5265** | **1196** | **17345** | **849** | **882740** |

**Supplementary Table 1:** Worldwide frequency distribution of different clades of SARS-CoV-2

| Worldwide | G | GH | GR | GV | GRY/B.1.1.7 | L | O | S | V | B.1.351 | P.1 | B.1.429+B.1.427 | B.1.525 | Total |
|---|---|---|---|---|---|---|---|---|---|---|---|---|---|---|
| Dec, 2019 | 0 | 0 | 0 | 0 | 0 | 22 | 3 | 1 | 0 | 0 | 0 | 0 | 0 | 26 |
| Jan, 2020 | 16 | 4 | 7 | 6 | 0 | 263 | 153 | 195 | 5 | 0 | 0 | 0 | 0 | 649 |
| Feb, 2020 | 171 | 51 | 96 | 3 | 1 | 339 | 482 | 265 | 242 | 0 | 0 | 0 | 0 | 1650 |
| Mar, 2020 | 12029 | 13976 | 9402 | 12 | 0 | 3050 | 2082 | 5492 | 4168 | 0 | 0 | 0 | 0 | 50211 |
| Apr, 2020 | 12672 | 12142 | 14424 | 5 | 0 | 1377 | 1785 | 1973 | 1715 | 0 | 0 | 0 | 0 | 46093 |
| May, 2020 | 5225 | 8043 | 8963 | 7 | 4 | 165 | 984 | 658 | 135 | 0 | 0 | 0 | 0 | 24184 |
| Jun, 2020 | 5139 | 6795 | 9154 | 100 | 0 | 56 | 467 | 260 | 60 | 0 | 0 | 0 | 0 | 22031 |
| Jul, 2020 | 5082 | 4382 | 17784 | 165 | 0 | 34 | 375 | 112 | 22 | 0 | 0 | 4 | 0 | 27960 |
| Aug, 2020 | 4787 | 7104 | 18866 | 2025 | 0 | 13 | 277 | 112 | 9 | 0 | 0 | 0 | 0 | 33193 |
| Sep, 2020 | 4997 | 7201 | 12710 | 10674 | 6 | 6 | 435 | 64 | 5 | 0 | 0 | 8 | 0 | 36106 |
| Oct, 2020 | 6446 | 9624 | 12805 | 23376 | 70 | 3 | 532 | 22 | 1 | 44 | 0 | 7 | 0 | 52930 |
| Nov, 2020 | 10887 | 15428 | 15974 | 27053 | 2231 | 8 | 244 | 31 | 0 | 271 | 0 | 246 | 0 | 72373 |
| Dec, 2020 | 13874 | 20358 | 12971 | 31415 | 13140 | 5 | 598 | 206 | 3 | 743 | 94 | 1389 | 14 | 94810 |
| Jan, 2021 | 15476 | 30215 | 12964 | 26318 | 59471 | 39 | 1192 | 451 | 0 | 1667 | 271 | 5856 | 152 | 154072 |
| Feb, 2021 | 14382 | 36196 | 14218 | 13752 | 81867 | 17 | 2648 | 495 | 0 | 1625 | 475 | 7522 | 348 | 173545 |
| Mar, 2021 | 6405 | 11547 | 7136 | 3684 | 57653 | 3 | 2393 | 167 | 0 | 915 | 356 | 2313 | 335 | 92907 |
| **Total** | **117588** | **183066** | **167474** | **138595** | **214443** | **5400** | **14650** | **10504** | **6365** | **5265** | **1196** | **17345** | **849** | **882740** |

**Supplementary Table 2:** Temporal distribution of different clades of SARS-CoV-2 globally

| No. | Asian country | G | GH | GR | GV | GRY/B.1.1.7 | L | O | S | V | B.1.351 | P.1 | B.1.429+B.1.427 | B.1.525 | Predominant Clade |
|---|---|---|---|---|---|---|---|---|---|---|---|---|---|---|---|
| 1 | Afghanistan | 3 | 13 | 0 | 0 | 0 | 0 | 0 | 0 | 0 | 0 | 0 | 0 | 0 | GH |
| 2 | Armenia | 0 | 0 | 13 | 0 | 0 | 0 | 2 | 0 | 0 | 0 | 0 | 0 | 0 | GR |
| 3 | Bahrain | 2 | 41 | 81 | 0 | 0 | 0 | 24 | 2 | 1 | 0 | 0 | 0 | 0 | GR |
| 4 | Bangladesh | 91 | 56 | 758 | 0 | 3 | 1 | 56 | 5 | 0 | 1 | 0 | 0 | 0 | GR |
| 5 | Brunei | 0 | 0 | 4 | 0 | 0 | 0 | 5 | 0 | 0 | 1 | 0 | 0 | 0 | S |
| 6 | Cambodia | 5 | 9 | 3 | 0 | 7 | 3 | 11 | 18 | 2 | 0 | 0 | 2 | 0 | O |
| 7 | China | 46 | 189 | 317 | 7 | 5 | 498 | 260 | 296 | 10 | 1 | 0 | 0 | 0 | L |
| 8 | Georgia | 3 | 9 | 6 | 0 | 2 | 0 | 4 | 1 | 3 | 0 | 0 | 0 | 0 | GH |
| 9 | Hong Kong | 300 | 375 | 314 | 19 | 8 | 41 | 167 | 10 | 20 | 0 | 0 | 0 | 0 | GH |
| 10 | India | 1254 | 1342 | 2324 | 5 | 81 | 26 | 928 | 111 | 7 | 4 | 0 | 0 | 0 | GR |
| 11 | Indonesia | 37 | 528 | 155 | 0 | 7 | 61 | 40 | 5 | 0 | 0 | 0 | 0 | 0 | GH |
| 12 | Iran | 43 | 14 | 6 | 1 | 1 | 4 | 214 | 0 | 0 | 0 | 0 | 0 | 0 | O |
| 13 | Iraq | 4 | 31 | 0 | 0 | 0 | 0 | 76 | 0 | 0 | 0 | 0 | 0 | 0 | O |
| 14 | Israel | 283 | 715 | 1339 | 110 | 1101 | 12 | 32 | 13 | 14 | 52 | 0 | 5 | 0 | GR |
| 15 | Japan | 220 | 452 | 20642 | 16 | 88 | 158 | 155 | 198 | 8 | 13 | 8 | 16 | 11 | GR |
| 16 | Jordan | 9 | 75 | 449 | 0 | 44 | 0 | 2 | 5 | 10 | 0 | 0 | 0 | 2 | GR |
| 17 | Kazakhstan | 8 | 6 | 22 | 0 | 0 | 1 | 29 | 19 | 0 | 0 | 0 | 0 | 0 | O |
| 18 | Kuwait | 8 | 12 | 1 | 0 | 1 | 0 | 10 | 0 | 0 | 0 | 0 | 0 | 0 | GH |
| 19 | Lebanon | 27 | 12 | 5 | 0 | 2 | 0 | 3 | 0 | 0 | 0 | 0 | 0 | 0 | G |
| 20 | Malaysia | 208 | 52 | 36 | 1 | 3 | 31 | 100 | 3 | 2 | 4 | 0 | 0 | 2 | G |
| 21 | Mongolia | 5 | 0 | 0 | 0 | 0 | 0 | 2 | 0 | 0 | 0 | 0 | 0 | 0 | G |
| 22 | Myanmar | 2 | 31 | 3 | 0 | 0 | 0 | 5 | 0 | 0 | 0 | 0 | 0 | 0 | GH |
| 23 | Nepal | 0 | 7 | 7 | 0 | 0 | 1 | 0 | 0 | 0 | 0 | 0 | 0 | 0 | GH/GR |
| 24 | Oman | 21 | 12 | 142 | 0 | 1 | 5 | 17 | 5 | 2 | 0 | 0 | 0 | 0 | GR |
| 25 | Pakistan | 40 | 37 | 6 | 1 | 3 | 3 | 15 | 3 | 0 | 0 | 0 | 0 | 0 | G |
| 26 | Palestine | 6 | 10 | 78 | 0 | 0 | 0 | 2 | 0 | 2 | 0 | 0 | 0 | 0 | GR |
| 27 | Philippines | 25 | 3 | 94 | 0 | 39 | 2 | 25 | 0 | 0 | 0 | 0 | 0 | 0 | GR |
| 28 | Qatar | 0 | 0 | 0 | 0 | 0 | 0 | 16 | 0 | 0 | 0 | 0 | 0 | 0 | O |
| 29 | Saudi Arabia | 59 | 522 | 80 | 0 | 0 | 0 | 254 | 38 | 0 | 0 | 0 | 0 | 0 | GH |
| 30 | Singapore | 141 | 257 | 243 | 12 | 69 | 114 | 984 | 39 | 23 | 13 | 0 | 3 | 2 | O |

| | | | | | | | | | | | | | | | |
|---|---|---|---|---|---|---|---|---|---|---|---|---|---|---|---|
| 31 | South Korea | 69 | 2145 | 214 | 16 | 102 | 5 | 15 | 63 | 155 | 5 | 1 | 47 | 1 | **GH** |
| 32 | Sri Lanka | 51 | 109 | 4 | 1 | 18 | 0 | 48 | 2 | 0 | 1 | 0 | 0 | 0 | **GH** |
| 33 | Taiwan | 19 | 51 | 25 | 1 | 4 | 15 | 29 | 11 | 11 | 0 | 0 | 7 | 0 | **GH** |
| 34 | Thailand | 40 | 351 | 67 | 4 | 9 | 15 | 171 | 239 | 7 | 4 | 0 | 0 | 1 | **GH** |
| 35 | Timor-Leste | 0 | 0 | 0 | 0 | 0 | 0 | 18 | 0 | 1 | 0 | 0 | 0 | 0 | **O** |
| 36 | United Arab Emirates | 117 | 182 | 1033 | 24 | 21 | 36 | 148 | 279 | 1 | 5 | 0 | 0 | 0 | **GR** |
| 37 | Uzbekistan | 0 | 0 | 0 | 0 | 0 | 0 | 2 | 0 | 0 | 0 | 0 | 0 | 0 | **O** |
| 38 | Vietnam | 9 | 33 | 73 | 1 | 11 | 7 | 2 | 12 | 5 | 0 | 0 | 0 | 0 | **GR** |
| | **Total** | **3155** | **7681** | **28544** | **219** | **1630** | **1039** | **3871** | **1377** | **284** | **104** | **9** | **80** | **19** | **48012** |

| No. | African country | G | GH | GR | GV | GRY/B.1.1.7 | L | O | S | V | B.1.351 | P.1 | B.1.429+B.1.427 | B.1.525 | Predominant Clade |
|---|---|---|---|---|---|---|---|---|---|---|---|---|---|---|---|
| 1 | Algeria | 10 | 9 | 4 | 0 | 0 | 0 | 6 | 0 | 0 | 0 | 0 | 0 | 0 | G |
| 2 | Benin | 2 | 4 | 0 | 0 | 0 | 0 | 3 | 3 | 0 | 0 | 0 | 0 | 0 | GH |
| 3 | Botswana | 1 | 6 | 7 | 0 | 0 | 0 | 0 | 1 | 0 | 53 | 0 | 0 | 0 | B.1.351 |
| 4 | Burkina Faso | 5 | 7 | 20 | 0 | 0 | 0 | 0 | 62 | 0 | 0 | 0 | 0 | 0 | S |
| 5 | Cameroon | 17 | 5 | 16 | 2 | 0 | 0 | 0 | 0 | 0 | 1 | 0 | 0 | 1 | G |
| 6 | Cote dIvoire | 7 | 16 | 2 | 0 | 0 | 0 | 0 | 40 | 0 | 0 | 0 | 0 | 0 | S |
| 7 | Democratic Republic of the Cong | 296 | 22 | 39 | 0 | 2 | 0 | 2 | 7 | 2 | 1 | 0 | 0 | 0 | G |
| 8 | Egypt | 57 | 150 | 160 | 0 | 0 | 16 | 53 | 11 | 0 | 0 | 0 | 0 | 0 | GR |
| 9 | Equatorial Guinea | 114 | 2 | 1 | 0 | 0 | 0 | 0 | 2 | 0 | 0 | 0 | 0 | 0 | G |
| 10 | Eswatini | 4 | 0 | 1 | 0 | 0 | 0 | 0 | 0 | 0 | 17 | 0 | 0 | 0 | B.1.351 |
| 11 | Ethiopia | 0 | 0 | 0 | 0 | 0 | 0 | 4 | 0 | 0 | 0 | 0 | 0 | 0 | O |
| 12 | Gabon | 1 | 1 | 0 | 0 | 0 | 0 | 5 | 2 | 0 | 0 | 0 | 0 | 0 | O |
| 13 | Gambia | 332 | 20 | 122 | 0 | 3 | 0 | 1 | 0 | 0 | 0 | 0 | 0 | 0 | G |
| 14 | Ghana | 19 | 47 | 169 | 4 | 116 | 0 | 11 | 32 | 0 | 4 | 0 | 0 | 6 | GR |
| 15 | Guinea | 6 | 1 | 0 | 0 | 0 | 0 | 0 | 1 | 0 | 0 | 0 | 0 | 0 | G |
| 16 | Kenya | 454 | 94 | 78 | 0 | 1 | 1 | 20 | 29 | 3 | 6 | 0 | 0 | 0 | G |
| 17 | Lesotho | 4 | 0 | 1 | 0 | 0 | 0 | 0 | 0 | 0 | 13 | 0 | 0 | 0 | B.1.351 |
| 18 | Libya | 0 | 0 | 0 | 0 | 0 | 0 | 1 | 0 | 0 | 0 | 0 | 0 | 0 | O |
| 19 | Madagascar | 0 | 2 | 2 | 0 | 0 | 0 | 2 | 0 | 0 | 0 | 0 | 0 | 0 | GH/GR/O |
| 20 | Mali | 2 | 5 | 0 | 0 | 0 | 0 | 3 | 14 | 0 | 0 | 0 | 0 | 0 | S |
| 21 | Martinique | 0 | 2 | 4 | 0 | 2 | 0 | 0 | 0 | 0 | 0 | 0 | 0 | 0 | GR |
| 22 | Mauritius | 0 | 5 | 1 | 2 | 1 | 0 | 0 | 1 | 0 | 2 | 0 | 0 | 0 | GH |
| 23 | Mayotte | 43 | 268 | 7 | 11 | 1 | 0 | 0 | 12 | 0 | 378 | 0 | 0 | 1 | B.1.351 |
| 24 | Morocco | 68 | 42 | 43 | 6 | 1 | 0 | 2 | 0 | 0 | 0 | 0 | 0 | 0 | G |
| 25 | Mozambique | 14 | 9 | 69 | 0 | 0 | 0 | 3 | 0 | 0 | 58 | 0 | 0 | 0 | GR |
| 26 | Namibia | 7 | 0 | 40 | 0 | 0 | 0 | 4 | 0 | 0 | 7 | 0 | 0 | 0 | GR |
| 27 | Nigeria | 46 | 62 | 203 | 1 | 125 | 0 | 4 | 29 | 9 | 0 | 0 | 0 | 103 | GR |
| 28 | Republic of the Congo | 19 | 20 | 2 | 0 | 0 | 0 | 12 | 0 | 0 | 0 | 0 | 0 | 0 | GH |
| 29 | Reunion | 5 | 32 | 0 | 8 | 0 | 0 | 0 | 0 | 0 | 16 | 0 | 0 | 0 | GH |
| 30 | Rwanda | 154 | 9 | 8 | 1 | 3 | 0 | 1 | 93 | 0 | 11 | 0 | 0 | 5 | G |
| 31 | Senegal | 137 | 26 | 44 | 0 | 1 | 25 | 33 | 4 | 0 | 0 | 0 | 0 | 0 | G |
| 32 | Sierra Leone | 1 | 2 | 2 | 0 | 0 | 0 | 1 | 5 | 0 | 0 | 0 | 0 | 0 | S |

| | | | | | | | | | | | | | | | |
|---|---|---|---|---|---|---|---|---|---|---|---|---|---|---|---|
| 33 | South Africa | 785 | 44 | 2019 | 14 | 1 | 4 | 31 | 5 | 5 | 1424 | 0 | 0 | 0 | **GR** |
| 34 | Togo | 0 | 2 | 1 | 0 | 0 | 0 | 1 | 3 | 0 | 0 | 0 | 0 | 0 | **S** |
| 35 | Tunisia | 5 | 42 | 17 | 19 | 1 | 0 | 10 | 2 | 0 | 0 | 0 | 0 | 0 | **GH** |
| 36 | Uganda | 45 | 6 | 14 | 0 | 0 | 0 | 19 | 121 | 3 | 0 | 0 | 0 | 0 | **S** |
| 37 | Union of the Comoros | 0 | 0 | 0 | 0 | 0 | 0 | 0 | 0 | 0 | 6 | 0 | 0 | 0 | **B.1.351** |
| 38 | Zambia | 8 | 6 | 161 | 0 | 0 | 0 | 11 | 4 | 0 | 31 | 0 | 0 | 0 | **GR** |
| 39 | Zimbabwe | 48 | 27 | 120 | 0 | 0 | 0 | 4 | 7 | 0 | 194 | 0 | 0 | 0 | **B.1.351** |
| | **Total** | **2716** | **995** | **3377** | **68** | **258** | **46** | **247** | **490** | **22** | **2222** | **0** | **0** | **116** | **10557** |

| No. | North America | G | GH | GR | GV | GRY/B.1.1.7 | L | O | S | V | B.1.351 | P.1 | B.1.429+B.1.427 | B.1.525 | Predominant clade |
|---|---|---|---|---|---|---|---|---|---|---|---|---|---|---|---|
| 1 | Antigua and Barbuda | 0 | 1 | 0 | 0 | 0 | 0 | 0 | 0 | 0 | 0 | 0 | 0 | 0 | GH |
| 2 | Barbados | 1 | 1 | 0 | 0 | 3 | 0 | 0 | 0 | 0 | 0 | 0 | 0 | 0 | GRY/B.1.1.7 |
| 3 | Belize | 0 | 2 | 2 | 0 | 0 | 0 | 0 | 0 | 0 | 0 | 0 | 0 | 0 | GH/GR |
| 4 | Bermuda | 8 | 18 | 0 | 4 | 0 | 0 | 0 | 0 | 0 | 0 | 0 | 0 | 0 | GH |
| 5 | British Virgin Islands | 0 | 2 | 1 | 1 | 0 | 0 | 0 | 0 | 0 | 0 | 0 | 1 | 0 | GH |
| 6 | Canada | 3072 | 11332 | 5995 | 31 | 612 | 30 | 80 | 506 | 127 | 20 | 37 | 13 | 11 | GH |
| 7 | Cayman Islands | 1 | 0 | 0 | 0 | 2 | 0 | 0 | 0 | 0 | 0 | 0 | 0 | 0 | GRY/B.1.1.7 |
| 8 | Costa Rica | 56 | 65 | 136 | 0 | 2 | 0 | 0 | 24 | 0 | 2 | 0 | 3 | 1 | GR |
| 9 | Cuba | 2 | 0 | 0 | 0 | 0 | 0 | 0 | 0 | 0 | 0 | 0 | 0 | 0 | G |
| 10 | Dominican Republic | 1 | 3 | 0 | 0 | 1 | 0 | 4 | 0 | 0 | 0 | 0 | 0 | 0 | O |
| 11 | El Salvador | 0 | 6 | 0 | 0 | 0 | 0 | 0 | 0 | 0 | 0 | 0 | 0 | 0 | GH |
| 12 | Guadeloupe | 13 | 23 | 6 | 3 | 6 | 0 | 0 | 0 | 0 | 1 | 0 | 3 | 0 | GH |
| 13 | Guatemala | 9 | 14 | 2 | 0 | 0 | 0 | 4 | 1 | 0 | 0 | 0 | 0 | 0 | GH |
| 14 | Jamaica | 0 | 3 | 0 | 1 | 4 | 0 | 1 | 0 | 4 | 0 | 0 | 0 | 0 | GH |
| 15 | Mexico | 946 | 234 | 1980 | 4 | 22 | 2 | 38 | 20 | 0 | 0 | 3 | 105 | 0 | GR |
| 16 | Panama | 141 | 28 | 3 | 1 | 0 | 0 | 7 | 388 | 2 | 1 | 0 | 0 | 0 | S |
| 17 | Saint Barthelemy | 1 | 1 | 0 | 0 | 0 | 0 | 0 | 0 | 0 | 0 | 0 | 0 | 0 | G/GH |
| 18 | Saint Kitts and Nevis | 0 | 3 | 0 | 0 | 0 | 0 | 0 | 0 | 0 | 0 | 0 | 0 | 0 | GH |
| 19 | Saint Lucia | 0 | 1 | 0 | 0 | 9 | 0 | 0 | 0 | 0 | 0 | 0 | 0 | 0 | GRY/B.1.1.7 |
| 20 | Saint Martin | 12 | 42 | 18 | 1 | 27 | 0 | 0 | 0 | 0 | 0 | 1 | 13 | 0 | GH |
| 21 | Saint Vincent and the Grenadin | 0 | 1 | 0 | 0 | 0 | 0 | 0 | 0 | 0 | 0 | 0 | 0 | 0 | GH |
| 22 | USA | 39384 | 116335 | 22121 | 213 | 6938 | 662 | 1948 | 3740 | 407 | 159 | 61 | 17006 | 111 | GH |
| | **Total** | **43647** | **128115** | **30264** | **259** | **7626** | **694** | **2082** | **4679** | **540** | **183** | **102** | **17144** | **123** | **235458** |

| No. | South America | G | GH | GR | GV | GRY/B.1.1.7 | L | O | S | V | B.1.351 | P.1 | B.1.429+B.1.427 | B.1.525 | Predominant clade |
|---|---|---|---|---|---|---|---|---|---|---|---|---|---|---|---|
| 1 | Argentina | 46 | 385 | 215 | 0 | 1 | 1 | 9 | 1 | 3 | 0 | 0 | 0 | 0 | GH |
| 2 | Aruba | 29 | 103 | 88 | 16 | 65 | 0 | 1 | 0 | 2 | 2 | 0 | 16 | 0 | GH |
| 3 | Bolivia | 20 | 0 | 17 | 0 | 0 | 0 | 0 | 1 | 0 | 0 | 0 | 0 | 0 | G |
| 4 | Bonaire | 0 | 10 | 0 | 2 | 56 | 0 | 0 | 0 | 0 | 0 | 0 | 0 | 0 | GRY/B.1.1.7 |
| 5 | Brazil | 282 | 79 | 3513 | 4 | 54 | 4 | 139 | 8 | 15 | 0 | 440 | 0 | 0 | GR |
| 6 | Chile | 192 | 197 | 997 | 0 | 27 | 2 | 3 | 20 | 7 | 0 | 6 | 7 | 0 | GR |
| 7 | Colombia | 159 | 266 | 111 | 1 | 0 | 2 | 2 | 10 | 4 | 0 | 23 | 1 | 0 | GH |
| 8 | Curacao | 15 | 32 | 21 | 5 | 46 | 6 | 0 | 0 | 0 | 0 | 0 | 0 | 0 | GRY/B.1.1.7 |
| 9 | Ecuador | 51 | 12 | 205 | 0 | 7 | 3 | 10 | 2 | 0 | 0 | 0 | 0 | 0 | GR |
| 10 | French Guiana | 0 | 87 | 53 | 4 | 3 | 0 | 0 | 1 | 0 | 0 | 4 | 0 | 0 | GH |
| 11 | Guatemala | 2 | 2 | 0 | 0 | 0 | 0 | 0 | 0 | 0 | 0 | 0 | 0 | 0 | G/GH |
| 12 | Paraguay | 7 | 1 | 7 | 0 | 0 | 0 | 0 | 0 | 0 | 0 | 0 | 0 | 0 | G |
| 13 | Peru | 103 | 31 | 781 | 3 | 3 | 2 | 5 | 22 | 1 | 0 | 23 | 0 | 0 | GR |
| 14 | Sint Eustatius | 0 | 4 | 0 | 0 | 0 | 0 | 0 | 0 | 0 | 0 | 0 | 0 | 0 | GH |
| 15 | Suriname | 66 | 0 | 5 | 0 | 0 | 0 | 0 | 0 | 0 | 0 | 0 | 0 | 0 | G |
| 16 | Trinidad | 0 | 16 | 0 | 0 | 1 | 0 | 0 | 0 | 0 | 0 | 0 | 0 | 0 | GH |
| 17 | Uruguay | 30 | 1 | 73 | 0 | 0 | 0 | 0 | 28 | 3 | 0 | 0 | 0 | 0 | GR |
| 18 | Venezuela | 9 | 2 | 1 | 0 | 0 | 0 | 0 | 0 | 0 | 0 | 0 | 0 | 0 | G |
| | **Total** | **1011** | **1228** | **6087** | **35** | **263** | **20** | **169** | **93** | **35** | **2** | **496** | **24** | **0** | **9463** |

| No. | Europe | G | GH | GR | GV | GRY/B.1.1.7 | L | O | S | V | B.1.351 | P.1 | B.1.429+B.1.427 | B.1.525 | Predominant clade |
|---|---|---|---|---|---|---|---|---|---|---|---|---|---|---|---|
| 1 | Albania | 1 | 1 | 4 | 0 | 24 | 0 | 0 | 0 | 0 | 0 | 0 | 0 | 0 | GRY/B.1.1.7 |
| 2 | Andorra | 0 | 0 | 0 | 0 | 0 | 0 | 1 | 0 | 0 | 0 | 0 | 0 | 0 | O |
| 3 | Austria | 771 | 520 | 617 | 177 | 400 | 28 | 24 | 16 | 17 | 166 | 0 | 2 | 3 | G |
| 4 | Azerbaijan | 0 | 0 | 7 | 0 | 0 | 0 | 2 | 0 | 0 | 0 | 0 | 0 | 0 | GR |
| 5 | Belarus | 11 | 16 | 10 | 0 | 1 | 0 | 2 | 0 | 0 | 0 | 0 | 0 | 0 | GH |
| 6 | Belgium | 2517 | 1353 | 1422 | 976 | 2760 | 68 | 404 | 42 | 70 | 425 | 87 | 1 | 11 | GRY/B.1.1.7 |
| 7 | Bosnia and Herzegovina | 41 | 13 | 11 | 1 | 21 | 0 | 2 | 0 | 1 | 0 | 0 | 0 | 0 | G |
| 8 | Bulgaria | 122 | 44 | 30 | 21 | 342 | 1 | 0 | 0 | 0 | 0 | 0 | 0 | 0 | GRY/B.1.1.7 |
| 9 | Croatia | 299 | 56 | 93 | 128 | 182 | 0 | 92 | 0 | 0 | 6 | 0 | 0 | 0 | G |
| 10 | Cyprus | 81 | 18 | 15 | 9 | 10 | 0 | 0 | 0 | 0 | 0 | 0 | 0 | 0 | G |
| 11 | Czech Republic | 418 | 62 | 192 | 12 | 343 | 9 | 194 | 3 | 4 | 1 | 0 | 0 | 0 | G |
| 12 | Denmark | 9237 | 7389 | 7189 | 21478 | 4889 | 49 | 31 | 100 | 5 | 12 | 0 | 25 | 121 | GV |
| 13 | Estonia | 117 | 15 | 137 | 123 | 97 | 0 | 0 | 0 | 0 | 2 | 0 | 0 | 0 | GR |
| 14 | Faroe Islands | 14 | 10 | 13 | 0 | 0 | 0 | 0 | 0 | 0 | 0 | 1 | 0 | 0 | G |
| 15 | Finland | 262 | 1289 | 311 | 61 | 393 | 12 | 22 | 3 | 10 | 6 | 0 | 1 | 4 | GH |
| 16 | France | 1695 | 4214 | 738 | 948 | 3894 | 29 | 951 | 228 | 32 | 387 | 17 | 4 | 12 | GH |
| 17 | Germany | 6209 | 3007 | 6080 | 7210 | 13728 | 231 | 836 | 212 | 66 | 505 | 55 | 5 | 86 | GRY/B.1.1.7 |
| 18 | Gibraltar | 33 | 40 | 68 | 493 | 128 | 0 | 0 | 0 | 0 | 0 | 0 | 0 | 0 | GV |
| 19 | Greece | 17 | 21 | 97 | 0 | 4 | 5 | 1 | 6 | 15 | 0 | 0 | 0 | 0 | GR |
| 20 | Hungary | 54 | 181 | 117 | 0 | 21 | 1 | 32 | 0 | 0 | 0 | 0 | 0 | 0 | GH |
| 21 | Iceland | 510 | 176 | 616 | 2675 | 20 | 36 | 29 | 17 | 93 | 0 | 0 | 0 | 0 | GV |
| 22 | Ireland | 490 | 191 | 776 | 983 | 3366 | 16 | 24 | 6 | 84 | 28 | 11 | 0 | 16 | GRY/B.1.1.7 |
| 23 | Italy | 2744 | 677 | 1793 | 4386 | 4478 | 3 | 380 | 87 | 121 | 17 | 319 | 1 | 47 | GV |
| 24 | Kosovo | 10 | 0 | 15 | 2 | 3 | 0 | 0 | 0 | 0 | 0 | 0 | 0 | 0 | GR |
| 25 | Latvia | 35 | 46 | 397 | 354 | 130 | 4 | 367 | 4 | 0 | 0 | 0 | 0 | 0 | GR |
| 26 | Liechtenstein | 36 | 2 | 0 | 0 | 1 | 0 |  | 0 | 0 | 0 | 0 | 0 | 0 | G |
| 27 | Lithuania | 189 | 14 | 398 | 1132 | 107 | 61 | 25 | 0 | 3 | 1 | 0 | 0 | 0 | GV |
| 28 | Luxembourg | 872 | 1333 | 277 | 689 | 32 | 5 | 0 | 8 | 7 | 2 | 0 | 0 | 0 | GH |
| 29 | Malta | 4 | 10 | 0 | 0 | 0 | 0 | 1 | 0 | 0 | 0 | 0 | 0 | 0 | GH |
| 30 | Moldova | 5 | 0 | 3 | 0 | 0 | 0 | 0 | 0 | 1 | 0 | 0 | 0 | 0 | G |
| 31 | Monaco | 0 | 2 | 1 | 2 | 1 | 0 | 0 | 0 | 0 | 1 | 0 | 0 | 0 | GV |

| | | | | | | | | | | | | | | | |
|---|---|---|---|---|---|---|---|---|---|---|---|---|---|---|---|
| 32 | Montenegro | 9 | 2 | 13 | 0 | 7 | 0 | 0 | 0 | 0 | 0 | 0 | 0 | 0 | GR |
| 33 | Netherlands | 4127 | 2113 | 1160 | 3975 | 3648 | 309 | 90 | 40 | 89 | 237 | 13 | 3 | 18 | G |
| 34 | North Macedonia | 80 | 15 | 235 | 12 | 53 | 0 | 0 | 0 | 0 | 0 | 0 | 1 | 0 | GR |
| 35 | Norway | 477 | 759 | 1350 | 579 | 1174 | 2 | 117 | 2 | 11 | 154 | 1 | 1 | 18 | GR |
| 36 | Poland | 603 | 90 | 568 | 92 | 824 | 1 | 6 | 1 | 16 | 4 | 0 | 0 | 0 | GRY/B.1.1.7 |
| 37 | Portugal | 717 | 387 | 1181 | 1104 | 755 | 18 | 19 | 51 | 86 | 12 | 11 | 0 | 0 | GR |
| 38 | Romania | 190 | 32 | 53 | 16 | 112 | 0 | 3 | 0 | 0 | 0 | 2 | 0 | 0 | G |
| 39 | Russia | 105 | 49 | 1898 | 1 | 4 | 2 | 33 | 2 | 1 | 3 | 0 | 0 | 0 | GR |
| 40 | Serbia | 23 | 23 | 168 | 4 | 2 | 0 | 1 | 0 | 0 | 0 | 0 | 0 | 0 | GR |
| 41 | Slovakia | 119 | 41 | 46 | 6 | 287 | 1 | 2 | 0 | 0 | 6 | 0 | 0 | 0 | GRY/B.1.1.7 |
| 42 | Slovenia | 3233 | 161 | 254 | 44 | 396 | 0 | 4 | 23 | 23 | 20 | 1 | 0 | 14 | G |
| 43 | Spain | 3053 | 566 | 1157 | 4591 | 2916 | 20 | 564 | 1400 | 135 | 22 | 6 | 2 | 7 | GV |
| 44 | Sweden | 1895 | 1130 | 929 | 2994 | 2078 | 29 | 381 | 12 | 1 | 172 | 11 | 2 | 1 | GV |
| 45 | Switzerland | 4549 | 5333 | 3200 | 4524 | 3169 | 13 | 364 | 26 | 32 | 102 | 22 | 4 | 5 | GH |
| 46 | Turkey | 569 | 486 | 1068 | 31 | 506 | 3 | 261 | 28 | 0 | 95 | 5 | 2 | 12 | GR |
| 47 | Ukraine | 42 | 0 | 75 | 3 | 22 | 0 | 1 | 0 | 1 | 0 | 0 | 0 | 0 | GR |
| 48 | United Kingdom | 19251 | 11453 | 50519 | 78101 | 153079 | 2574 | 2503 | 656 | 4230 | 316 | 22 | 15 | 210 | GRY/B.1.1.7 |
| | **Total** | **65836** | **43340** | **85301** | **137937** | **204407** | **3530** | **7769** | **2973** | **5154** | **2702** | **584** | **69** | **585** | **560187** |

| No. | Oceania | G | GH | GR | GV | GRY/B.1.1.7 | L | O | S | V | B.1.351 | P.1 | B.1.429+B.1.427 | B.1.525 | Predominant clade |
|---|---|---|---|---|---|---|---|---|---|---|---|---|---|---|---|
| 1 | Australia | 954 | 1407 | 13516 | 40 | 177 | 46 | 416 | 838 | 253 | 32 | 2 | 16 | 6 | GR |
| 2 | Guam | 9 | 8 | 2 | 0 | 0 | 0 | 4 | 0 | 0 | 0 | 0 | 7 | 0 | G |
| 3 | New Zealand | 240 | 263 | 375 | 36 | 82 | 25 | 33 | 53 | 77 | 20 | 3 | 4 | 0 | GR |
| 4 | Papua New Guinea | 0 | 7 | 0 | 0 | 0 | 0 | 59 | 0 | 0 | 0 | 0 | 0 | 0 | GH |
| 5 | Northern Mariana Islands | 20 | 22 | 8 | 1 | 0 | 0 | 0 | 1 | 0 | 0 | 0 | 1 | 0 | O |
| | **Total** | **1223** | **1707** | **13901** | **77** | **259** | **71** | **512** | **892** | **330** | **52** | **5** | **28** | **6** | **19063** |

**Supplementary Table 3:** Frequency distribution of different clades of SARS-CoV-2 among countries of six different continents

| Asia | G | GH | GR | GV | GRY/B.1.1.7 | L | O | S | V | B.1.351 | P.1 | B.1.429+B.1.427 | B.1.525 | Total |
|---|---|---|---|---|---|---|---|---|---|---|---|---|---|---|
| Dec, 2019 | 0 | 0 | 0 | 0 | 0 | 21 | 2 | 1 | 0 | 0 | 0 | 0 | 0 | 24 |
| Jan, 2020 | 5 | 1 | 1 | 0 | 0 | 254 | 123 | 190 | 2 | 0 | 0 | 0 | 0 | 576 |
| Feb, 2020 | 5 | 17 | 4 | 0 | 0 | 288 | 404 | 207 | 116 | 0 | 0 | 0 | 0 | 1041 |
| Mar, 2020 | 317 | 481 | 619 | 0 | 0 | 342 | 696 | 492 | 139 | 0 | 0 | 0 | 0 | 3086 |
| Apr, 2020 | 333 | 486 | 2927 | 1 | 0 | 75 | 1013 | 85 | 24 | 0 | 0 | 0 | 0 | 4944 |
| May, 2020 | 481 | 456 | 1772 | 0 | 0 | 30 | 786 | 278 | 1 | 0 | 0 | 0 | 0 | 3804 |
| Jun, 2020 | 320 | 828 | 1456 | 0 | 0 | 16 | 374 | 68 | 1 | 0 | 0 | 0 | 0 | 3063 |
| Jul, 2020 | 314 | 428 | 3442 | 0 | 0 | 10 | 154 | 10 | 1 | 0 | 0 | 0 | 0 | 4359 |
| Aug, 2020 | 182 | 537 | 4343 | 2 | 0 | 3 | 102 | 5 | 0 | 0 | 0 | 0 | 0 | 5174 |
| Sep, 2020 | 127 | 400 | 1762 | 13 | 0 | 0 | 35 | 3 | 0 | 0 | 0 | 0 | 0 | 2340 |
| Oct, 2020 | 105 | 438 | 1788 | 40 | 0 | 0 | 20 | 1 | 0 | 0 | 0 | 0 | 0 | 2392 |
| Nov, 2020 | 145 | 827 | 3024 | 22 | 2 | 0 | 48 | 8 | 0 | 0 | 0 | 0 | 0 | 4076 |
| Dec, 2020 | 308 | 1144 | 4250 | 69 | 243 | 0 | 20 | 14 | 0 | 13 | 0 | 16 | 0 | 6077 |
| Jan, 2021 | 298 | 911 | 2756 | 54 | 723 | 0 | 48 | 8 | 0 | 31 | 7 | 42 | 11 | 4889 |
| Feb, 2021 | 183 | 615 | 350 | 18 | 648 | 0 | 39 | 7 | 0 | 43 | 2 | 22 | 6 | 1933 |
| Mar, 2021 | 32 | 112 | 50 | 0 | 14 | 0 | 7 | 0 | 0 | 17 | 0 | 0 | 2 | 234 |
| **Total** | **3155** | **7681** | **28544** | **219** | **1630** | **1039** | **3871** | **1377** | **284** | **104** | **9** | **80** | **19** | **48012** |

| Africa | G | GH | GR | GV | GRY/B.1.1.7 | L | O | S | V | B.1.351 | P.1 | B.1.429+B.1.427 | B.1.525 | Total |
|---|---|---|---|---|---|---|---|---|---|---|---|---|---|---|
| Dec, 2019 | 0 | 0 | 0 | 0 | 0 | 0 | 0 | 0 | 0 | 0 | 0 | 0 | 0 | 0 |
| Jan, 2020 | 0 | 0 | 0 | 0 | 0 | 0 | 0 | 0 | 0 | 0 | 0 | 0 | 0 | 0 |
| Feb, 2020 | 1 | 1 | 1 | 0 | 0 | 0 | 0 | 0 | 1 | 0 | 0 | 0 | 0 | 4 |
| Mar, 2020 | 108 | 64 | 48 | 3 | 0 | 4 | 52 | 42 | 15 | 0 | 0 | 0 | 0 | 336 |
| Apr, 2020 | 358 | 78 | 122 | 0 | 0 | 2 | 32 | 37 | 3 | 0 | 0 | 0 | 0 | 632 |
| May, 2020 | 408 | 81 | 234 | 0 | 0 | 0 | 13 | 42 | 3 | 0 | 0 | 0 | 0 | 781 |
| Jun, 2020 | 297 | 79 | 433 | 0 | 0 | 5 | 26 | 33 | 0 | 0 | 0 | 0 | 0 | 873 |
| Jul, 2020 | 386 | 59 | 965 | 1 | 0 | 10 | 16 | 26 | 0 | 0 | 0 | 0 | 0 | 1463 |
| Aug, 2020 | 258 | 92 | 347 | 0 | 0 | 1 | 9 | 55 | 0 | 0 | 0 | 0 | 0 | 762 |
| Sep, 2020 | 109 | 27 | 277 | 7 | 0 | 0 | 2 | 52 | 0 | 0 | 0 | 0 | 0 | 474 |
| Oct, 2020 | 158 | 32 | 270 | 7 | 0 | 0 | 1 | 12 | 0 | 40 | 0 | 0 | 0 | 520 |
| Nov, 2020 | 147 | 74 | 206 | 10 | 0 | 0 | 8 | 2 | 0 | 291 | 0 | 0 | 0 | 738 |
| Dec, 2020 | 157 | 96 | 225 | 9 | 12 | 2 | 28 | 85 | 0 | 740 | 0 | 0 | 6 | 1360 |
| Jan, 2021 | 268 | 312 | 217 | 30 | 224 | 20 | 47 | 82 | 0 | 1001 | 0 | 0 | 58 | 2259 |
| Feb, 2021 | 59 | 0 | 32 | 1 | 22 | 2 | 11 | 22 | 0 | 143 | 0 | 0 | 36 | 328 |
| Mar, 2021 | 2 | 0 | 0 | 0 | 0 | 0 | 2 | 0 | 0 | 7 | 0 | 0 | 16 | 27 |
| **Total** | **2716** | **995** | **3377** | **68** | **258** | **46** | **247** | **490** | **22** | **2222** | **0** | **0** | **116** | **10557** |

| North America | G | GH | GR | GV | GRY/B.1.1.7 | L | O | S | V | B.1.351 | P.1 | B.1.429+B.1.427 | B.1.525 | Total |
|---|---|---|---|---|---|---|---|---|---|---|---|---|---|---|
| Dec, 2019 | 0 | 0 | 0 | 0 | 0 | 0 | 0 | 0 | 0 | 0 | 0 | 0 | 0 | 0 |
| Jan, 2020 | 1 | 0 | 0 | 0 | 0 | 8 | 3 | 7 | 0 | 0 | 0 | 0 | 0 | 19 |
| Feb, 2020 | 1 | 6 | 9 | 0 | 0 | 14 | 38 | 68 | 4 | 0 | 0 | 0 | 0 | 140 |
| Mar, 2020 | 1828 | 8473 | 1108 | 0 | 0 | 354 | 308 | 2756 | 402 | 0 | 0 | 1 | 0 | 15230 |
| Apr, 2020 | 2401 | 7729 | 1473 | 0 | 0 | 268 | 125 | 1093 | 126 | 0 | 0 | 0 | 0 | 13216 |
| May, 2020 | 1704 | 5638 | 1468 | 2 | 4 | 31 | 81 | 287 | 7 | 0 | 0 | 0 | 0 | 9222 |
| Jun, 2020 | 2872 | 4798 | 1593 | 1 | 0 | 5 | 41 | 125 | 0 | 0 | 0 | 0 | 0 | 9435 |
| Jul, 2020 | 2724 | 3180 | 2197 | 1 | 0 | 0 | 128 | 19 | 0 | 0 | 0 | 4 | 0 | 8254 |
| Aug, 2020 | 2865 | 3632 | 1756 | 6 | 0 | 3 | 53 | 11 | 1 | 0 | 0 | 0 | 0 | 8327 |
| Sep, 2020 | 2246 | 3246 | 1602 | 10 | 0 | 1 | 67 | 2 | 0 | 0 | 0 | 8 | 0 | 7182 |
| Oct, 2020 | 3435 | 4745 | 2095 | 28 | 0 | 0 | 27 | 4 | 0 | 0 | 0 | 8 | 0 | 10342 |
| Nov, 2020 | 4510 | 9593 | 2565 | 15 | 2 | 5 | 96 | 11 | 0 | 0 | 0 | 267 | 0 | 17064 |
| Dec, 2020 | 5320 | 14757 | 2604 | 35 | 115 | 2 | 297 | 16 | 0 | 1 | 0 | 1306 | 1 | 24454 |
| Jan, 2021 | 5845 | 21536 | 2849 | 101 | 1216 | 3 | 376 | 111 | 0 | 26 | 12 | 5761 | 30 | 37866 |
| Feb, 2021 | 6021 | 30764 | 6178 | 56 | 4393 | 0 | 222 | 118 | 0 | 108 | 42 | 7364 | 50 | 55315 |
| Mar, 2021 | 1873 | 10018 | 2767 | 4 | 1896 | 0 | 220 | 51 | 0 | 48 | 48 | 2425 | 42 | 19392 |
| **Total** | **43647** | **128115** | **30264** | **259** | **7626** | **694** | **2082** | **4679** | **540** | **183** | **102** | **17144** | **123** | **235458** |

| South America | G | GH | GR | GV | GRY/B.1.1.7 | L | O | S | V | B.1.351 | P.1 | B.1.429+B.1.427 | B.1.525 | Total |
|---|---|---|---|---|---|---|---|---|---|---|---|---|---|---|
| Dec, 2019 | 0 | 0 | 0 | 0 | 0 | 0 | 0 | 0 | 0 | 0 | 0 | 0 | 0 | 0 |
| Jan, 2020 | 0 | 0 | 0 | 0 | 0 | 0 | 0 | 0 | 0 | 0 | 0 | 0 | 0 | 0 |
| Feb, 2020 | 3 | 0 | 3 | 0 | 0 | 0 | 0 | 0 | 2 | 0 | 0 | 0 | 0 | 8 |
| Mar, 2020 | 137 | 172 | 538 | 0 | 0 | 15 | 52 | 62 | 22 | 0 | 0 | 0 | 0 | 998 |
| Apr, 2020 | 152 | 163 | 846 | 0 | 0 | 3 | 9 | 5 | 8 | 0 | 0 | 0 | 0 | 1186 |
| May, 2020 | 70 | 177 | 744 | 0 | 0 | 0 | 7 | 0 | 3 | 0 | 0 | 0 | 0 | 1001 |
| Jun, 2020 | 54 | 72 | 483 | 0 | 0 | 1 | 5 | 1 | 0 | 0 | 0 | 0 | 0 | 616 |
| Jul, 2020 | 38 | 95 | 489 | 0 | 0 | 0 | 12 | 1 | 0 | 0 | 0 | 0 | 0 | 635 |
| Aug, 2020 | 38 | 135 | 423 | 0 | 0 | 0 | 0 | 22 | 0 | 0 | 0 | 0 | 0 | 618 |
| Sep, 2020 | 7 | 35 | 334 | 0 | 0 | 0 | 30 | 0 | 0 | 0 | 0 | 0 | 0 | 406 |
| Oct, 2020 | 13 | 58 | 385 | 1 | 0 | 0 | 45 | 0 | 0 | 0 | 0 | 0 | 0 | 502 |
| Nov, 2020 | 16 | 33 | 457 | 0 | 0 | 1 | 3 | 0 | 0 | 0 | 0 | 0 | 0 | 510 |
| Dec, 2020 | 132 | 73 | 483 | 2 | 11 | 0 | 1 | 1 | 0 | 0 | 89 | 1 | 0 | 793 |
| Jan, 2021 | 92 | 123 | 483 | 17 | 52 | 0 | 3 | 0 | 0 | 0 | 248 | 4 | 0 | 1022 |
| Feb, 2021 | 113 | 70 | 363 | 15 | 102 | 0 | 1 | 1 | 0 | 2 | 142 | 12 | 0 | 821 |
| Mar, 2021 | 146 | 22 | 56 | 0 | 98 | 0 | 1 | 0 | 0 | 0 | 17 | 7 | 0 | 347 |
| **Total** | **1011** | **1228** | **6087** | **35** | **263** | **20** | **169** | **93** | **35** | **2** | **496** | **24** | **0** | **9463** |

| Europe | G | GH | GR | GV | GRY/B.1.1.7 | L | O | S | V | B.1.351 | P.1 | B.1.429+B.1.427 | B.1.525 | Total |
|---|---|---|---|---|---|---|---|---|---|---|---|---|---|---|
| Dec, 2019 | 0 | 0 | 0 | 0 | 0 | 0 | 1 | 0 | 0 | 0 | 0 | 0 | 0 | 1 |
| Jan, 2020 | 9 | 3 | 5 | 6 | 0 | 4 | 13 | 3 | 3 | 0 | 0 | 0 | 0 | 46 |
| Feb, 2020 | 183 | 27 | 88 | 3 | 1 | 39 | 61 | 16 | 117 | 0 | 0 | 0 | 0 | 535 |
| Mar, 2020 | 9246 | 4089 | 7414 | 7 | 0 | 2275 | 710 | 1547 | 3294 | 0 | 0 | 0 | 0 | 28582 |
| Apr, 2020 | 9220 | 2932 | 9914 | 4 | 0 | 1020 | 536 | 483 | 1522 | 0 | 0 | 0 | 0 | 25631 |
| May, 2020 | 2740 | 1287 | 5418 | 5 | 0 | 102 | 131 | 85 | 121 | 0 | 0 | 0 | 0 | 9889 |
| Jun, 2020 | 1440 | 761 | 5328 | 99 | 0 | 29 | 63 | 35 | 59 | 0 | 0 | 0 | 0 | 7814 |
| Jul, 2020 | 714 | 443 | 4059 | 176 | 0 | 13 | 46 | 55 | 21 | 0 | 0 | 0 | 0 | 5527 |
| Aug, 2020 | 1768 | 2118 | 6885 | 2171 | 0 | 6 | 108 | 11 | 8 | 0 | 0 | 0 | 0 | 13075 |
| Sep, 2020 | 2952 | 3042 | 7509 | 9740 | 6 | 3 | 273 | 4 | 5 | 0 | 0 | 0 | 0 | 23534 |
| Oct, 2020 | 2120 | 4130 | 7958 | 23031 | 63 | 3 | 433 | 5 | 1 | 0 | 0 | 0 | 0 | 37744 |
| Nov, 2020 | 5992 | 6015 | 9071 | 27141 | 1974 | 2 | 80 | 10 | 0 | 1 | 0 | 0 | 0 | 50286 |
| Dec, 2020 | 8101 | 4653 | 5170 | 32522 | 15779 | 1 | 205 | 82 | 3 | 68 | 0 | 3 | 3 | 66590 |
| Jan, 2021 | 9319 | 7946 | 5794 | 26510 | 57207 | 15 | 721 | 226 | 0 | 542 | 27 | 24 | 82 | 108413 |
| Feb, 2021 | 8233 | 4831 | 6919 | 13635 | 74398 | 15 | 2292 | 309 | 0 | 1266 | 310 | 30 | 272 | 112510 |
| Mar, 2021 | 3799 | 1063 | 3769 | 2887 | 54979 | 3 | 2096 | 102 | 0 | 825 | 247 | 12 | 228 | 70010 |
| **Total** | **65836** | **43340** | **85301** | **137937** | **204407** | **3530** | **7769** | **2973** | **5154** | **2702** | **584** | **69** | **585** | **560187** |

| Oceania | G | GH | GR | GV | GRY/B.1.1.7 | L | O | S | V | B.1.351 | P.1 | B.1.429+B.1.427 | B.1.525 | Total |
|---|---|---|---|---|---|---|---|---|---|---|---|---|---|---|
| Dec, 2019 | 0 | 0 | 0 | 0 | 0 | 0 | 0 | 0 | 0 | 0 | 0 | 0 | 0 | 0 |
| Jan, 2020 | 1 | 0 | 0 | 0 | 0 | 1 | 14 | 5 | 0 | 0 | 0 | 0 | 0 | 21 |
| Feb, 2020 | 0 | 0 | 0 | 0 | 0 | 2 | 13 | 4 | 0 | 0 | 0 | 0 | 0 | 19 |
| Mar, 2020 | 526 | 484 | 364 | 2 | 0 | 60 | 262 | 597 | 292 | 0 | 0 | 0 | 0 | 2587 |
| Apr, 2020 | 344 | 231 | 148 | 0 | 0 | 6 | 101 | 257 | 38 | 0 | 0 | 0 | 0 | 1125 |
| May, 2020 | 24 | 178 | 41 | 0 | 0 | 0 | 30 | 7 | 0 | 0 | 0 | 0 | 0 | 280 |
| Jun, 2020 | 13 | 61 | 507 | 0 | 0 | 0 | 8 | 4 | 0 | 0 | 0 | 0 | 0 | 593 |
| Jul, 2020 | 123 | 47 | 6414 | 0 | 0 | 0 | 9 | 1 | 0 | 0 | 0 | 0 | 0 | 6594 |
| Aug, 2020 | 34 | 24 | 4942 | 2 | 0 | 0 | 5 | 2 | 0 | 0 | 0 | 0 | 0 | 5009 |
| Sep, 2020 | 24 | 34 | 1063 | 2 | 0 | 2 | 7 | 2 | 0 | 0 | 0 | 0 | 0 | 1134 |
| Oct, 2020 | 47 | 62 | 248 | 24 | 0 | 0 | 6 | 0 | 0 | 0 | 0 | 0 | 0 | 387 |
| Nov, 2020 | 25 | 98 | 39 | 21 | 1 | 0 | 2 | 0 | 0 | 0 | 0 | 0 | 0 | 186 |
| Dec, 2020 | 30 | 210 | 44 | 16 | 26 | 0 | 40 | 4 | 0 | 8 | 0 | 9 | 0 | 387 |
| Jan, 2021 | 24 | 100 | 47 | 9 | 94 | 0 | 6 | 4 | 0 | 26 | 0 | 14 | 2 | 326 |
| Feb, 2021 | 2 | 106 | 18 | 0 | 62 | 0 | 2 | 1 | 0 | 6 | 0 | 5 | 2 | 204 |
| Mar, 2021 | 6 | 72 | 26 | 1 | 76 | 0 | 7 | 4 | 0 | 12 | 5 | 0 | 2 | 211 |
| **Total** | **1223** | **1707** | **13901** | **77** | **259** | **71** | **512** | **892** | **330** | **52** | **5** | **28** | **6** | **19063** |

**Supplementary Table 4:** Temporal distribution of different clades of SARS-CoV-2 in six different continents

| No. | Country Name | Continent | Total confirmed cases | Total confirmed deaths | CFR |
|---|---|---|---|---|---|
| 1 | Mexico | North America | 2226550 | 201623 | 9.06 |
| 2 | Egypt | Africa | 200739 | 11914 | 5.94 |
| 3 | Ecuador | South America | 325124 | 16746 | 5.15 |
| 4 | China | Asia | 102715 | 4851 | 4.72 |
| 5 | Bolivia | South America | 269628 | 12186 | 4.52 |
| 6 | Afghanistan | Asia | 56384 | 2476 | 4.39 |
| 7 | Zimbabwe | Africa | 36839 | 1520 | 4.13 |
| 8 | Union of the Comoros | Africa | 3696 | 146 | 3.95 |
| 9 | Bosnia and Herzegovina | Europe | 165787 | 6427 | 3.88 |
| 10 | Bulgaria | Europe | 333250 | 12913 | 3.87 |
| 11 | Mali | Africa | 9921 | 384 | 3.87 |
| 12 | Eswatini | Africa | 17326 | 667 | 3.85 |
| 13 | Guatemala | North America | 193556 | 6809 | 3.52 |
| 14 | Guatemala | South America | 193556 | 6809 | 3.52 |
| 15 | Tunisia | Africa | 251169 | 8760 | 3.49 |
| 16 | South Africa | Africa | 1545979 | 52710 | 3.41 |
| 17 | Peru | South America | 1529882 | 51469 | 3.36 |
| 18 | Iran | Asia | 1864984 | 62478 | 3.35 |
| 19 | Hungary | Europe | 645733 | 20435 | 3.16 |
| 20 | El Salvador | North America | 64431 | 2003 | 3.11 |
| 21 | Greece | Europe | 255755 | 7945 | 3.11 |
| 22 | Australia | Oceania | 29278 | 909 | 3.10 |
| 23 | Italy | Europe | 3544957 | 108350 | 3.06 |
| 24 | Gambia | Africa | 5420 | 164 | 3.03 |
| 25 | Lesotho | Africa | 10686 | 315 | 2.95 |
| 26 | North Macedonia | Europe | 127240 | 3716 | 2.92 |
| 27 | United Kingdom | Europe | 4341736 | 126670 | 2.92 |
| 28 | Germany | Europe | 2791822 | 76093 | 2.73 |
| 29 | Senegal | Africa | 38566 | 1044 | 2.71 |
| 30 | Indonesia | Asia | 1501093 | 40581 | 2.70 |
| 31 | Slovakia | Europe | 359330 | 9624 | 2.68 |
| 32 | Colombia | South America | 2382730 | 62955 | 2.64 |
| 33 | Democratic Republic of the Cong | Africa | 28011 | 739 | 2.64 |
| 34 | Algeria | Africa | 116946 | 3084 | 2.64 |
| 35 | Belgium | Europe | 872936 | 22921 | 2.63 |
| 36 | Republic of the Congo | Africa | 27794 | 726 | 2.61 |
| 37 | Belize | North America | 12411 | 317 | 2.55 |
| 38 | Brazil | South America | 12534688 | 312206 | 2.49 |
| 39 | Romania | Europe | 940443 | 23234 | 2.47 |
| 40 | Antigua and Barbuda | North America | 1136 | 28 | 2.46 |
| 41 | Argentina | South America | 2308597 | 55449 | 2.40 |
| 42 | Canada | North America | 965404 | 22880 | 2.37 |
| 43 | Chile | South America | 984484 | 23070 | 2.34 |
| 44 | Spain | Europe | 3270825 | 75199 | 2.30 |
| 45 | Poland | Europe | 2288826 | 52392 | 2.29 |
| 46 | Myanmar | Asia | 142393 | 3206 | 2.25 |
| 47 | Croatia | Europe | 269009 | 5928 | 2.20 |

| | | | | | |
|---|---|---|---|---|---|
| 48 | Gibraltar | Europe | 4273 | 94 | 2.20 |
| 49 | Russia | Europe | 4536820 | 98442 | 2.17 |
| 50 | Pakistan | Asia | 659116 | 14256 | 2.16 |
| 51 | Moldova | Europe | 228370 | 4915 | 2.15 |
| 52 | France | Europe | 4481165 | 94402 | 2.11 |
| 53 | Kosovo | Europe | 88712 | 1834 | 2.07 |
| 54 | Portugal | Europe | 820716 | 16843 | 2.05 |
| 55 | Slovenia | Europe | 212965 | 4311 | 2.02 |
| 56 | Sierra Leone | Africa | 3970 | 79 | 1.99 |
| 57 | Ireland | Europe | 235078 | 4667 | 1.99 |
| 58 | Liechtenstein | Europe | 2747 | 54 | 1.97 |
| 59 | Ukraine | Europe | 1662942 | 32418 | 1.95 |
| 60 | Paraguay | South America | 208655 | 4063 | 1.95 |
| 61 | Suriname | South America | 9097 | 177 | 1.95 |
| 62 | Japan | Asia | 470175 | 9086 | 1.93 |
| 63 | Latvia | Europe | 101207 | 1883 | 1.86 |
| 64 | Armenia | Asia | 191491 | 3497 | 1.83 |
| 65 | USA | North America | 29968464 | 544430 | 1.82 |
| 66 | Philippines | Asia | 731894 | 13186 | 1.80 |
| 67 | Hong Kong | Asia | 11,454 | 205 | 1.79 |
| 68 | Albania | Europe | 124419 | 2216 | 1.78 |
| 69 | Trinidad | South America | 7977 | 142 | 1.78 |
| 70 | Morocco | Africa | 494756 | 8807 | 1.78 |
| 71 | Guam | Oceania | 7594 | 134 | 1.76 |
| 72 | Panama | North America | 353839 | 6100 | 1.72 |
| 73 | Czech Republic | Europe | 1523668 | 26222 | 1.72 |
| 74 | Sweden | Europe | 780018 | 13402 | 1.72 |
| 75 | Saudi Arabia | Asia | 388866 | 6656 | 1.71 |
| 76 | Iraq | Asia | 838265 | 14249 | 1.70 |
| 77 | Libya | Africa | 157545 | 2653 | 1.68 |
| 78 | Austria | Europe | 536751 | 9030 | 1.68 |
| 79 | South Korea | Asia | 103000 | 1729 | 1.68 |
| 80 | Lithuania | Europe | 215216 | 3566 | 1.66 |
| 81 | Madagascar | Africa | 23969 | 394 | 1.64 |
| 82 | Kenya | Africa | 131116 | 2135 | 1.63 |
| 83 | Switzerland | Europe | 594043 | 9589 | 1.61 |
| 84 | Jamaica | North America | 38514 | 586 | 1.52 |
| 85 | Cameroon | Africa | 47669 | 721 | 1.51 |
| 86 | Guadeloupe | North America | 11512 | 173 | 1.50 |
| 87 | Bangladesh | Asia | 600895 | 8949 | 1.49 |
| 88 | Equatorial Guinea | Africa | 6914 | 102 | 1.48 |
| 89 | Brunei | Asia | 207 | 3 | 1.45 |
| 90 | Rwanda | Africa | 21490 | 305 | 1.42 |
| 91 | Ethiopia | Africa | 202545 | 2825 | 1.39 |
| 92 | Saint Lucia | North America | 4202 | 58 | 1.38 |
| 93 | Montenegro | Europe | 90416 | 1245 | 1.38 |
| 94 | Azerbaijan | Europe | 257330 | 3513 | 1.37 |
| 95 | Zambia | Africa | 88012 | 1200 | 1.36 |
| 96 | Costa Rica | North America | 215178 | 2931 | 1.36 |

| | | | | | |
|---|---|---|---|---|---|
| 97 | Vietnam | Asia | 2594 | 35 | 1.35 |
| 98 | Georgia | Asia | 281145 | 3773 | 1.34 |
| 99 | Malta | Europe | 28938 | 388 | 1.34 |
| 100 | India | Asia | 12095855 | 162114 | 1.34 |
| 101 | Lebanon | Asia | 462339 | 6136 | 1.33 |
| 102 | Botswana | Africa | 38466 | 506 | 1.32 |
| 103 | Dominican Republic | North America | 252182 | 3307 | 1.31 |
| 104 | Netherlands | Europe | 1259155 | 16475 | 1.31 |
| 105 | Benin | Africa | 7100 | 90 | 1.27 |
| 106 | Nigeria | Africa | 162641 | 2049 | 1.26 |
| 107 | Kazakhstan | Asia | 293761 | 3696 | 1.26 |
| 108 | Northern Mariana Islands | Oceania | 159 | 2 | 1.26 |
| 109 | Monaco | Europe | 2265 | 28 | 1.24 |
| 110 | New Zealand | Oceania | 2139 | 26 | 1.22 |
| 111 | Luxembourg | Europe | 61073 | 741 | 1.21 |
| 112 | Namibia | Africa | 43923 | 513 | 1.17 |
| 113 | Bermuda | North America | 1028 | 12 | 1.17 |
| 114 | Mozambique | Africa | 67292 | 769 | 1.14 |
| 115 | Burkina Faso | Africa | 12702 | 145 | 1.14 |
| 116 | Barbados | North America | 3629 | 41 | 1.13 |
| 117 | Jordan | Asia | 597256 | 6651 | 1.11 |
| 118 | Nepal | Asia | 276980 | 3027 | 1.09 |
| 119 | Palestine | Asia | 240000 | 2614 | 1.09 |
| 120 | Finland | Europe | 76425 | 822 | 1.08 |
| 121 | Togo | Africa | 9992 | 107 | 1.07 |
| 122 | Oman | Asia | 156883 | 1662 | 1.06 |
| 123 | Denmark | Europe | 228692 | 2415 | 1.06 |
| 124 | Mauritius | Africa | 963 | 10 | 1.04 |
| 125 | Venezuela | South America | 156655 | 1565 | 1.00 |
| 126 | Papua New Guinea | Oceania | 5620 | 56 | 1.00 |
| 127 | Taiwan | Asia | 1023 | 10 | 0.98 |
| 128 | Andorra | Europe | 11888 | 115 | 0.97 |
| 129 | Turkey | Europe | 3240577 | 31230 | 0.96 |
| 130 | Uruguay | South America | 97406 | 915 | 0.94 |
| 131 | Serbia | Europe | 590018 | 5231 | 0.89 |
| 132 | Aruba | South America | 9259 | 82 | 0.89 |
| 133 | Estonia | Europe | 105416 | 896 | 0.85 |
| 134 | Uganda | Africa | 40820 | 335 | 0.82 |
| 135 | Ghana | Africa | 90287 | 740 | 0.82 |
| 136 | Mayotte | Africa | 19306 | 154 | 0.80 |
| 137 | Bonaire | South America | 1299 | 10 | 0.77 |
| 138 | Uzbekistan | Asia | 82682 | 628 | 0.76 |
| 139 | Israel | Asia | 831084 | 6165 | 0.74 |
| 140 | Saint Martin | North America | 1657 | 12 | 0.72 |
| 141 | Norway | Europe | 93145 | 660 | 0.71 |
| 142 | Belarus | Europe | 319599 | 2227 | 0.70 |
| 143 | Reunion | Africa | 15561 | 102 | 0.66 |
| 144 | Martinique | Africa | 7679 | 50 | 0.65 |
| 145 | British Virgin Islands | North America | 154 | 1 | 0.65 |

| | | | | | |
|---|---|---|---|---|---|
| **146** | Guinea | Africa | 19773 | 123 | 0.62 |
| **147** | Sri Lanka | Asia | 92303 | 566 | 0.61 |
| **148** | Gabon | Africa | 19140 | 114 | 0.60 |
| **149** | Saint Vincent and the Grenadin | North America | 1739 | 10 | 0.58 |
| **150** | Cuba | North America | 73204 | 417 | 0.57 |
| **151** | Kuwait | Asia | 229550 | 1298 | 0.57 |
| **152** | Cyprus | Europe | 44991 | 254 | 0.56 |
| **153** | Cote dIvoire | Africa | 43422 | 239 | 0.55 |
| **154** | French Guiana | South America | 16922 | 89 | 0.53 |
| **155** | Iceland | Europe | 6183 | 29 | 0.47 |
| **156** | Cambodia | Asia | 2378 | 11 | 0.46 |
| **157** | Cayman Islands | North America | 487 | 2 | 0.41 |
| **158** | Curacao | South America | 7558 | 30 | 0.40 |
| **159** | Malaysia | Asia | 342885 | 1260 | 0.37 |
| **160** | Bahrain | Asia | 142669 | 515 | 0.36 |
| **161** | Thailand | Asia | 28773 | 94 | 0.33 |
| **162** | United Arab Emirates | Asia | 457071 | 1486 | 0.33 |
| **163** | Qatar | Asia | 178464 | 286 | 0.16 |
| **164** | Faroe Islands | Europe | 661 | 1 | 0.15 |
| **165** | Saint Barthelemy | North America | 857 | 1 | 0.12 |
| **166** | Mongolia | Asia | 7589 | 6 | 0.08 |
| **167** | Singapore | Asia | 60321 | 30 | 0.05 |
| **168** | Timor-Leste | Asia | 512 | 0 | 0.00 |
| **169** | Saint Kitts and Nevis | North America | 44 | 0 | 0.00 |
| **170** | Sint Eustatius | South America | 20 | 0 | 0.00 |
| | | **Overall** | **126849230** | **2777747** | **2.19** |

**Supplementary Table 5:**Case Fatality Rate of 170 countries from six different continents

| Asian country | Confirmed cases | Confirmed deaths | CFR |
|---|---|---|---|
| China | 102715 | 4851 | 4.72 |
| Afghanistan | 56384 | 2476 | 4.39 |
| Iran | 1864984 | 62478 | 3.35 |
| Indonesia | 1501093 | 40581 | 2.70 |
| Myanmar | 142393 | 3206 | 2.25 |
| Pakistan | 659116 | 14256 | 2.16 |
| Japan | 470175 | 9086 | 1.93 |
| Armenia | 191491 | 3497 | 1.83 |
| Philippines | 731894 | 13186 | 1.80 |
| Hong Kong | 11,454 | 205 | 1.79 |
| Saudi Arabia | 388866 | 6656 | 1.71 |
| Iraq | 838265 | 14249 | 1.70 |
| South Korea | 103000 | 1729 | 1.68 |
| Bangladesh | 600895 | 8949 | 1.49 |
| Brunei | 207 | 3 | 1.45 |
| Vietnam | 2594 | 35 | 1.35 |
| Georgia | 281145 | 3773 | 1.34 |
| India | 12095855 | 162114 | 1.34 |
| Lebanon | 462339 | 6136 | 1.33 |
| Kazakhstan | 293761 | 3696 | 1.26 |
| Jordan | 597256 | 6651 | 1.11 |
| Nepal | 276980 | 3027 | 1.09 |
| Palestine | 240000 | 2614 | 1.09 |
| Oman | 156883 | 1662 | 1.06 |
| Taiwan | 1023 | 10 | 0.98 |
| Uzbekistan | 82682 | 628 | 0.76 |
| Israel | 831084 | 6165 | 0.74 |
| Sri Lanka | 92303 | 566 | 0.61 |
| Kuwait | 229550 | 1298 | 0.57 |
| Cambodia | 2378 | 11 | 0.46 |
| Malaysia | 342885 | 1260 | 0.37 |
| Bahrain | 142669 | 515 | 0.36 |
| Thailand | 28773 | 94 | 0.33 |
| United Arab Emirates | 457071 | 1486 | 0.33 |
| Qatar | 178464 | 286 | 0.16 |
| Mongolia | 7589 | 6 | 0.08 |
| Singapore | 60321 | 30 | 0.05 |
| Timor-Leste | 512 | 0 | 0.00 |
| **Overall** | **24527049** | **387471** | **1.58** |

| Africa country | Confirmed cases | Confirmed deaths | CFR |
|---|---|---|---|
| Egypt | 200739 | 11914 | 5.94 |
| Zimbabwe | 36839 | 1520 | 4.13 |
| Union of the Comoros | 3696 | 146 | 3.95 |
| Mali | 9921 | 384 | 3.87 |
| Eswatini | 17326 | 667 | 3.85 |
| Tunisia | 251169 | 8760 | 3.49 |
| South Africa | 1545979 | 52710 | 3.41 |
| Gambia | 5420 | 164 | 3.03 |
| Lesotho | 10686 | 315 | 2.95 |
| Senegal | 38566 | 1044 | 2.71 |
| Democratic Republic of the Cong | 28011 | 739 | 2.64 |
| Algeria | 116946 | 3084 | 2.64 |
| Republic of the Congo | 27794 | 726 | 2.61 |
| Sierra Leone | 3970 | 79 | 1.99 |
| Morocco | 494756 | 8807 | 1.78 |
| Libya | 157545 | 2653 | 1.68 |
| Madagascar | 23969 | 394 | 1.64 |
| Kenya | 131116 | 2135 | 1.63 |
| Cameroon | 47669 | 721 | 1.51 |
| Equatorial Guinea | 6914 | 102 | 1.48 |
| Rwanda | 21490 | 305 | 1.42 |
| Ethiopia | 202545 | 2825 | 1.39 |
| Zambia | 88012 | 1200 | 1.36 |
| Botswana | 38466 | 506 | 1.32 |
| Benin | 7100 | 90 | 1.27 |
| Nigeria | 162641 | 2049 | 1.26 |
| Namibia | 43923 | 513 | 1.17 |
| Mozambique | 67292 | 769 | 1.14 |
| Burkina Faso | 12702 | 145 | 1.14 |
| Togo | 9992 | 107 | 1.07 |
| Mauritius | 963 | 10 | 1.04 |
| Uganda | 40820 | 335 | 0.82 |
| Ghana | 90287 | 740 | 0.82 |
| Mayotte | 19306 | 154 | 0.80 |
| Reunion | 15561 | 102 | 0.66 |
| Martinique | 7679 | 50 | 0.65 |
| Guinea | 19773 | 123 | 0.62 |
| Gabon | 19140 | 114 | 0.60 |
| Cote dIvoire | 43422 | 239 | 0.55 |
| **Overall** | **4070145** | **107440** | **2.64** |

| Europe | Confirmed cases | Confirmed deaths | CFR |
|---|---|---|---|
| Bosnia and Herzegovina | 165787 | 6427 | 3.88 |
| Bulgaria | 333250 | 12913 | 3.87 |
| Hungary | 645733 | 20435 | 3.16 |
| Greece | 255755 | 7945 | 3.11 |
| Italy | 3544957 | 108350 | 3.06 |
| North Macedonia | 127240 | 3716 | 2.92 |
| United Kingdom | 4341736 | 126670 | 2.92 |
| Germany | 2791822 | 76093 | 2.73 |
| Slovakia | 359330 | 9624 | 2.68 |
| Belgium | 872936 | 22921 | 2.63 |
| Romania | 940443 | 23234 | 2.47 |
| Spain | 3270825 | 75199 | 2.30 |
| Poland | 2288826 | 52392 | 2.29 |
| Croatia | 269009 | 5928 | 2.20 |
| Gibraltar | 4273 | 94 | 2.20 |
| Russia | 4536820 | 98442 | 2.17 |
| Moldova | 228370 | 4915 | 2.15 |
| France | 4481165 | 94402 | 2.11 |
| Kosovo | 88712 | 1834 | 2.07 |
| Portugal | 820716 | 16843 | 2.05 |
| Slovenia | 212965 | 4311 | 2.02 |
| Ireland | 235078 | 4667 | 1.99 |
| Liechtenstein | 2747 | 54 | 1.97 |
| Ukraine | 1662942 | 32418 | 1.95 |
| Latvia | 101207 | 1883 | 1.86 |
| Albania | 124419 | 2216 | 1.78 |
| Czech Republic | 1523668 | 26222 | 1.72 |
| Sweden | 780018 | 13402 | 1.72 |
| Austria | 536751 | 9030 | 1.68 |
| Lithuania | 215216 | 3566 | 1.66 |
| Switzerland | 594043 | 9589 | 1.61 |
| Montenegro | 90416 | 1245 | 1.38 |
| Azerbaijan | 257330 | 3513 | 1.37 |
| Malta | 28938 | 388 | 1.34 |
| Netherlands | 1259155 | 16475 | 1.31 |
| Monaco | 2265 | 28 | 1.24 |
| Luxembourg | 61073 | 741 | 1.21 |
| Finland | 76425 | 822 | 1.08 |
| Denmark | 228692 | 2415 | 1.06 |
| Andorra | 11888 | 115 | 0.97 |
| Turkey | 3240577 | 31230 | 0.96 |
| Serbia | 590018 | 5231 | 0.89 |
| Estonia | 105416 | 896 | 0.85 |
| Norway | 93145 | 660 | 0.71 |
| Belarus | 319599 | 2227 | 0.70 |

| | | | |
|---|---|---|---|
| Cyprus | 44991 | 254 | 0.56 |
| Iceland | 6183 | 29 | 0.47 |
| Faroe Islands | 661 | 1 | 0.15 |
| **Overall** | **42773531** | **942005** | **2.20** |

| North America | Confirmed cases | Confirmed deaths | CFR |
|---|---|---|---|
| Mexico | 2226550 | 201623 | 9.06 |
| Guatemala | 193556 | 6809 | 3.52 |
| El Salvador | 64431 | 2003 | 3.11 |
| Belize | 12411 | 317 | 2.55 |
| Antigua and Barbuda | 1136 | 28 | 2.46 |
| Canada | 965404 | 22880 | 2.37 |
| USA | 29968464 | 544430 | 1.82 |
| Panama | 353839 | 6100 | 1.72 |
| Jamaica | 38514 | 586 | 1.52 |
| Guadeloupe | 11512 | 173 | 1.50 |
| Saint Lucia | 4202 | 58 | 1.38 |
| Costa Rica | 215178 | 2931 | 1.36 |
| Dominican Republic | 252182 | 3307 | 1.31 |
| Bermuda | 1028 | 12 | 1.17 |
| Barbados | 3629 | 41 | 1.13 |
| Saint Martin | 1657 | 12 | 0.72 |
| British Virgin Islands | 154 | 1 | 0.65 |
| Saint Vincent and the Grenadin | 1739 | 10 | 0.58 |
| Cuba | 73204 | 417 | 0.57 |
| Cayman Islands | 487 | 2 | 0.41 |
| Saint Barthelemy | 857 | 1 | 0.12 |
| Saint Kitts and Nevis | 44 | 0 | 0.00 |
| **Overall** | **34390178** | **791741** | **2.30** |

| Oceania | **Confirmed cases** | **Confirmed deaths** | **CFR** |
|---|---|---|---|
| Australia | 29278 | 909 | 3.10 |
| Guam | 7594 | 134 | 1.76 |
| Northern Mariana Islands | 159 | 2 | 1.26 |
| New Zealand | 2139 | 26 | 1.22 |
| Papua New Guinea | 5620 | 56 | 1.00 |
| **Overall** | **44790** | **1127** | **2.52** |

| South America | Confirmed cases | Confirmed deaths | CFR |
|---|---|---|---|
| Ecuador | 325124 | 16746 | 5.15 |
| Bolivia | 269628 | 12186 | 4.52 |
| Guatemala | 193556 | 6809 | 3.52 |
| Peru | 1529882 | 51469 | 3.36 |
| Colombia | 2382730 | 62955 | 2.64 |
| Brazil | 12534688 | 312206 | 2.49 |
| Argentina | 2308597 | 55449 | 2.40 |
| Chile | 984484 | 23070 | 2.34 |
| Paraguay | 208655 | 4063 | 1.95 |
| Suriname | 9097 | 177 | 1.95 |
| Trinidad | 7977 | 142 | 1.78 |
| Venezuela | 156655 | 1565 | 1.00 |
| Uruguay | 97406 | 915 | 0.94 |
| Aruba | 9259 | 82 | 0.89 |
| Bonaire | 1299 | 10 | 0.77 |
| French Guiana | 16922 | 89 | 0.53 |
| Curacao | 7558 | 30 | 0.40 |
| Sint Eustatius | 20 | 0 | 0.00 |
| **Overall** | **21043537** | **547963** | **2.60** |

**Supplementary Table 6:** Case Fatality Rate of 170 countries from six different continents

| No. | Country | Continent | Total confirmed cases | Total confirmed deaths | CFR | Predominant clade |
|---|---|---|---|---|---|---|
| 1 | Bolivia | South America | 269628 | 12186 | 4.52 | G |
| 2 | Bosnia and Herzegovina | Europe | 165787 | 6427 | 3.88 | G |
| 3 | Gambia | Africa | 5420 | 164 | 3.03 | G |
| 4 | Senegal | Africa | 38566 | 1044 | 2.71 | G |
| 5 | Democratic Republic of the Cong | Africa | 28011 | 739 | 2.64 | G |
| 6 | Algeria | Africa | 116946 | 3084 | 2.64 | G |
| 7 | Romania | Europe | 940443 | 23234 | 2.47 | G |
| 8 | Croatia | Europe | 269009 | 5928 | 2.20 | G |
| 9 | Pakistan | Asia | 659116 | 14256 | 2.16 | G |
| 10 | Moldova | Europe | 228370 | 4915 | 2.15 | G |
| 11 | Slovenia | Europe | 212965 | 4311 | 2.02 | G |
| 12 | Liechtenstein | Europe | 2747 | 54 | 1.97 | G |
| 13 | Paraguay | South America | 208655 | 4063 | 1.95 | G |
| 14 | Suriname | South America | 9097 | 177 | 1.95 | G |
| 15 | Morocco | Africa | 494756 | 8807 | 1.78 | G |
| 16 | Guam | Oceania | 7594 | 134 | 1.76 | G |
| 17 | Czech Republic | Europe | 1523668 | 26222 | 1.72 | G |
| 18 | Austria | Europe | 536751 | 9030 | 1.68 | G |
| 19 | Kenya | Africa | 131116 | 2135 | 1.63 | G |
| 20 | Cameroon | Africa | 47669 | 721 | 1.51 | G |
| 21 | Equatorial Guinea | Africa | 6914 | 102 | 1.48 | G |
| 22 | Rwanda | Africa | 21490 | 305 | 1.42 | G |
| 23 | Lebanon | Asia | 462339 | 6136 | 1.33 | G |
| 24 | Netherlands | Europe | 1259155 | 16475 | 1.31 | G |
| 25 | Venezuela | South America | 156655 | 1565 | 1.00 | G |
| 26 | Guinea | Africa | 19773 | 123 | 0.62 | G |
| 27 | Cuba | North America | 73204 | 417 | 0.57 | G |
| 28 | Cyprus | Europe | 44991 | 254 | 0.56 | G |
| 29 | Malaysia | Asia | 342885 | 1260 | 0.37 | G |
| 30 | Faroe Islands | Europe | 661 | 1 | 0.15 | G |
| 31 | Mongolia | Asia | 7589 | 6 | 0.08 | G |
| 32 | Afghanistan | Asia | 56384 | 2476 | 4.39 | GH |

| | | | | | | |
|---|---|---|---|---|---|---|
| 33 | Guatemala | North America | 193556 | 6809 | 3.52 | **GH** |
| 34 | Tunisia | Africa | 251169 | 8760 | 3.49 | **GH** |
| 35 | Hungary | Europe | 645733 | 20435 | 3.16 | **GH** |
| 36 | El Salvador | North America | 64431 | 2003 | 3.11 | **GH** |
| 37 | Indonesia | Asia | 1501093 | 40581 | 2.70 | **GH** |
| 38 | Colombia | South America | 2382730 | 62955 | 2.64 | **GH** |
| 39 | Republic of the Congo | Africa | 27794 | 726 | 2.61 | **GH** |
| 40 | Antigua and Barbuda | North America | 1136 | 28 | 2.46 | **GH** |
| 41 | Argentina | South America | 2308597 | 55449 | 2.40 | **GH** |
| 42 | Canada | North America | 965404 | 22880 | 2.37 | **GH** |
| 43 | Myanmar | Asia | 142393 | 3206 | 2.25 | **GH** |
| 44 | France | Europe | 4481165 | 94402 | 2.11 | **GH** |
| 45 | USA | North America | 29968464 | 544430 | 1.82 | **GH** |
| 46 | Hong Kong | Asia | 11,454 | 205 | 1.79 | **GH** |
| 47 | Trinidad | South America | 7977 | 142 | 1.78 | **GH** |
| 48 | Saudi Arabia | Asia | 388866 | 6656 | 1.71 | **GH** |
| 49 | South Korea | Asia | 103000 | 1729 | 1.68 | **GH** |
| 50 | Switzerland | Europe | 594043 | 9589 | 1.61 | **GH** |
| 51 | Jamaica | North America | 38514 | 586 | 1.52 | **GH** |
| 52 | Guadeloupe | North America | 11512 | 173 | 1.50 | **GH** |
| 53 | Georgia | Asia | 281145 | 3773 | 1.34 | **GH** |
| 54 | Malta | Europe | 28938 | 388 | 1.34 | **GH** |
| 55 | Benin | Africa | 7100 | 90 | 1.27 | **GH** |
| 56 | Northern Mariana Islands | Oceania | 159 | 2 | 1.26 | **GH** |
| 57 | Luxembourg | Europe | 61073 | 741 | 1.21 | **GH** |
| 58 | Bermuda | North America | 1028 | 12 | 1.17 | **GH** |
| 59 | Finland | Europe | 76425 | 822 | 1.08 | **GH** |
| 60 | Mauritius | Africa | 963 | 10 | 1.04 | **GH** |
| 61 | Taiwan | Asia | 1023 | 10 | 0.98 | **GH** |
| 62 | Aruba | South America | 9259 | 82 | 0.89 | **GH** |
| 63 | Saint Martin | North America | 1657 | 12 | 0.72 | **GH** |
| 64 | Belarus | Europe | 319599 | 2227 | 0.70 | **GH** |
| 65 | Reunion | Africa | 15561 | 102 | 0.66 | **GH** |

| | | | | | | |
|---|---|---|---|---|---|---|
| 66 | British Virgin Islands | North America | 154 | 1 | 0.65 | **GH** |
| 67 | Sri Lanka | Asia | 92303 | 566 | 0.61 | **GH** |
| 68 | Saint Vincent and the Grenadin | North America | 1739 | 10 | 0.58 | **GH** |
| 69 | Kuwait | Asia | 229550 | 1298 | 0.57 | **GH** |
| 70 | French Guiana | South America | 16922 | 89 | 0.53 | **GH** |
| 71 | Thailand | Asia | 28773 | 94 | 0.33 | **GH** |
| 72 | Saint Kitts and Nevis | North America | 44 | 0 | 0.00 | **GH** |
| 73 | Sint Eustatius | South America | 20 | 0 | 0.00 | **GH** |
| 74 | Mexico | North America | 2226550 | 201623 | 9.06 | **GR** |
| 75 | Egypt | Africa | 200739 | 11914 | 5.94 | **GR** |
| 76 | Ecuador | South America | 325124 | 16746 | 5.15 | **GR** |
| 77 | South Africa | Africa | 1545979 | 52710 | 3.41 | **GR** |
| 78 | Peru | South America | 1529882 | 51469 | 3.36 | **GR** |
| 79 | Greece | Europe | 255755 | 7945 | 3.11 | **GR** |
| 80 | Australia | Oceania | 29278 | 909 | 3.10 | **GR** |
| 81 | North Macedonia | Europe | 127240 | 3716 | 2.92 | **GR** |
| 82 | Brazil | South America | 12534688 | 312206 | 2.49 | **GR** |
| 83 | Chile | South America | 984484 | 23070 | 2.34 | **GR** |
| 84 | Russia | Europe | 4536820 | 98442 | 2.17 | **GR** |
| 85 | Kosovo | Europe | 88712 | 1834 | 2.07 | **GR** |
| 86 | Portugal | Europe | 820716 | 16843 | 2.05 | **GR** |
| 87 | Ukraine | Europe | 1662942 | 32418 | 1.95 | **GR** |
| 88 | Japan | Asia | 470175 | 9086 | 1.93 | **GR** |
| 89 | Latvia | Europe | 101207 | 1883 | 1.86 | **GR** |
| 90 | Armenia | Asia | 191491 | 3497 | 1.83 | **GR** |
| 91 | Philippines | Asia | 731894 | 13186 | 1.80 | **GR** |
| 92 | Bangladesh | Asia | 600895 | 8949 | 1.49 | **GR** |
| 93 | Montenegro | Europe | 90416 | 1245 | 1.38 | **GR** |
| 94 | Azerbaijan | Europe | 257330 | 3513 | 1.37 | **GR** |
| 95 | Zambia | Africa | 88012 | 1200 | 1.36 | **GR** |
| 96 | Costa Rica | North America | 215178 | 2931 | 1.36 | **GR** |
| 97 | Vietnam | Asia | 2594 | 35 | 1.35 | **GR** |
| 98 | India | Asia | 12095855 | 162114 | 1.34 | **GR** |

| | | | | | | |
|---|---|---|---|---|---|---|
| 99 | Nigeria | Africa | 162641 | 2049 | 1.26 | **GR** |
| 100 | New Zealand | Oceania | 2139 | 26 | 1.22 | **GR** |
| 101 | Namibia | Africa | 43923 | 513 | 1.17 | **GR** |
| 102 | Mozambique | Africa | 67292 | 769 | 1.14 | **GR** |
| 103 | Jordan | Asia | 597256 | 6651 | 1.11 | **GR** |
| 104 | Palestine | Asia | 240000 | 2614 | 1.09 | **GR** |
| 105 | Oman | Asia | 156883 | 1662 | 1.06 | **GR** |
| 106 | Turkey | Europe | 3240577 | 31230 | 0.96 | **GR** |
| 107 | Uruguay | South America | 97406 | 915 | 0.94 | **GR** |
| 108 | Serbia | Europe | 590018 | 5231 | 0.89 | **GR** |
| 109 | Estonia | Europe | 105416 | 896 | 0.85 | **GR** |
| 110 | Ghana | Africa | 90287 | 740 | 0.82 | **GR** |
| 111 | Israel | Asia | 831084 | 6165 | 0.74 | **GR** |
| 112 | Norway | Europe | 93145 | 660 | 0.71 | **GR** |
| 113 | Martinique | Africa | 7679 | 50 | 0.65 | **GR** |
| 114 | Bahrain | Asia | 142669 | 515 | 0.36 | **GR** |
| 115 | United Arab Emirates | Asia | 457071 | 1486 | 0.33 | **GR** |
| 116 | Italy | Europe | 3544957 | 108350 | 3.06 | **GV** |
| 117 | Spain | Europe | 3270825 | 75199 | 2.30 | **GV** |
| 118 | Gibraltar | Europe | 4273 | 94 | 2.20 | **GV** |
| 119 | Sweden | Europe | 780018 | 13402 | 1.72 | **GV** |
| 120 | Lithuania | Europe | 215216 | 3566 | 1.66 | **GV** |
| 121 | Monaco | Europe | 2265 | 28 | 1.24 | **GV** |
| 122 | Denmark | Europe | 228692 | 2415 | 1.06 | **GV** |
| 123 | Iceland | Europe | 6183 | 29 | 0.47 | **GV** |
| 124 | China | Asia | 102715 | 4851 | 4.72 | **L** |
| 125 | Mali | Africa | 9921 | 384 | 3.87 | **S** |
| 126 | Sierra Leone | Africa | 3970 | 79 | 1.99 | **S** |
| 127 | Panama | North America | 353839 | 6100 | 1.72 | **S** |
| 128 | Brunei | Asia | 207 | 3 | 1.45 | **S** |
| 129 | Burkina Faso | Africa | 12702 | 145 | 1.14 | **S** |
| 130 | Togo | Africa | 9992 | 107 | 1.07 | **S** |
| 131 | Uganda | Africa | 40820 | 335 | 0.82 | **S** |

| | | | | | | |
|---|---|---|---|---|---|---|
| 132 | Cote dIvoire | Africa | 43422 | 239 | 0.55 | **S** |
| 133 | Iran | Asia | 1864984 | 62478 | 3.35 | **O** |
| 134 | Iraq | Asia | 838265 | 14249 | 1.70 | **O** |
| 135 | Libya | Africa | 157545 | 2653 | 1.68 | **O** |
| 136 | Ethiopia | Africa | 202545 | 2825 | 1.39 | **O** |
| 137 | Dominican Republic | North America | 252182 | 3307 | 1.31 | **O** |
| 138 | Kazakhstan | Asia | 293761 | 3696 | 1.26 | **O** |
| 139 | Papua New Guinea | Oceania | 5620 | 56 | 1.00 | **O** |
| 140 | Andorra | Europe | 11888 | 115 | 0.97 | **O** |
| 141 | Uzbekistan | Asia | 82682 | 628 | 0.76 | **O** |
| 142 | Gabon | Africa | 19140 | 114 | 0.60 | **O** |
| 143 | Cambodia | Asia | 2378 | 11 | 0.46 | **O** |
| 144 | Qatar | Asia | 178464 | 286 | 0.16 | **O** |
| 145 | Singapore | Asia | 60321 | 30 | 0.05 | **O** |
| 146 | Timor-Leste | Asia | 512 | 0 | 0.00 | **O** |
| 147 | Guatemala | South America | 193556 | 6809 | 3.52 | **G/GH** |
| 148 | Saint Barthelemy | North America | 857 | 1 | 0.12 | **G/GH** |
| 149 | Belize | North America | 12411 | 317 | 2.55 | **GH/GR** |
| 150 | Nepal | Asia | 276980 | 3027 | 1.09 | **GH/GR** |
| 151 | Madagascar | Africa | 23969 | 394 | 1.64 | **GH/GR/O** |
| 152 | Bulgaria | Europe | 333250 | 12913 | 3.87 | **GRY/B.1.1.7** |
| 153 | United Kingdom | Europe | 4341736 | 126670 | 2.92 | **GRY/B.1.1.7** |
| 154 | Germany | Europe | 2791822 | 76093 | 2.73 | **GRY/B.1.1.7** |
| 155 | Slovakia | Europe | 359330 | 9624 | 2.68 | **GRY/B.1.1.7** |
| 156 | Belgium | Europe | 872936 | 22921 | 2.63 | **GRY/B.1.1.7** |
| 157 | Poland | Europe | 2288826 | 52392 | 2.29 | **GRY/B.1.1.7** |
| 158 | Ireland | Europe | 235078 | 4667 | 1.99 | **GRY/B.1.1.7** |
| 159 | Albania | Europe | 124419 | 2216 | 1.78 | **GRY/B.1.1.7** |
| 160 | Saint Lucia | North America | 4202 | 58 | 1.38 | **GRY/B.1.1.7** |
| 161 | Barbados | North America | 3629 | 41 | 1.13 | **GRY/B.1.1.7** |
| 162 | Bonaire | South America | 1299 | 10 | 0.77 | **GRY/B.1.1.7** |
| 163 | Cayman Islands | North America | 487 | 2 | 0.41 | **GRY/B.1.1.7** |
| 164 | Curacao | South America | 7558 | 30 | 0.40 | **GRY/B.1.1.7** |

| | | | | | | |
|---|---|---|---|---|---|---|
| 165 | Zimbabwe | Africa | 36839 | 1520 | 4.13 | **B.1.351** |
| 166 | Union of the Comoros | Africa | 3696 | 146 | 3.95 | **B.1.351** |
| 167 | Eswatini | Africa | 17326 | 667 | 3.85 | **B.1.351** |
| 168 | Lesotho | Africa | 10686 | 315 | 2.95 | **B.1.351** |
| 169 | Botswana | Africa | 38466 | 506 | 1.32 | **B.1.351** |
| 170 | Mayotte | Africa | 19306 | 154 | 0.80 | **B.1.351** |

**Supplementary Table 7:** Case Fatality Rate of different countries according to predominant clade
